# Miro1 Mediates Skeletal Muscle Insulin Resistance in Type 2 Diabetes

**DOI:** 10.1101/2025.08.06.25333143

**Authors:** Elizabeth C. Heintz, Wagner S. Dantas, Analisa L. Taylor, Elizabeth R. M. Zunica, Kathryn P. Belmont, Jacob T. Mey, Robbie Beyl, Bolormaa Vandanmagsar, Hailey A. Parry, Peter Ajayi, Brian Glancy, Christopher L. Axelrod, John P. Kirwan

## Abstract

Imbalanced skeletal muscle mitochondrial dynamics contributed to the onset and progression of type 2 diabetes (T2D) by mechanisms that remained incompletely understood. Here, we examined the role of mitochondrial Rho GTPase 1 (Miro1), an outer mitochondrial membrane enzyme, in the regulation of skeletal muscle insulin action and glucose homeostasis in T2D. Miro1 accumulated in the skeletal muscle of mice and humans with obesity and T2D, a phenomenon driven by impaired insulin-mediated interaction between AKT and Miro1 at the outer mitochondrial membrane. To determine whether Miro1 accumulation was reversible and functionally linked to metabolic improvements, we prospectively evaluated the impact of exercise training on skeletal muscle Miro1 expression, mitochondrial function, and insulin sensitivity in patients with T2D. Patients with T2D (N=24) were randomized to 12 weeks of standard care or exercise training. At baseline and after 12 weeks, we assessed changes in whole-body metabolic and mitochondrial function. Exercise training reduced skeletal muscle Miro1 accumulation (64.3% vs. –53.2% change from baseline; p=0.001) and enhanced mitochondrial oxidative capacity (–37.7% vs. 216.3% change from baseline; p=0.005) and insulin sensitivity (–18.5% vs. 80.0% change from baseline; p=0.007). To further establish a causal role for Miro1 in glucose homeostasis, we generated muscle-specific Miro1 loss-of-function models in mice and cells. Muscle-specific deletion of Miro1 improved insulin action and oxidative capacity in both models. Taken together, these findings supported a key regulatory role for skeletal muscle Miro1 in the pathophysiology of T2D.

## INTRODUCTION

Type 2 diabetes (T2D) has emerged as one of the major public health and economic burdens of the 21st century, affecting more than 37 million people in the United States and more than 500 million people globally (∼10.5%).^1^ As these shocking numbers continue to rise, the cost of caring for patients with diabetes and its complications is placing enormous strain on the global economy, totaling nearly $1 trillion dollars in economic impact,^2^ despite the development of nearly a dozen new medications for disease management. As the scale of the problem grows exponentially, our fundamental understanding of the cellular and molecular mechanisms of T2D remains incomplete, restricting our ability to develop effective and targeted medications that treat the underlying pathophysiology rather than symptoms of disease. T2D arises when insulin secretion is insufficient to compensate for progressive and sustained peripheral insulin resistance, causing a systemic rise in glucose concentrations. Skeletal muscle insulin resistance is a primary and rate-limiting defect in the onset and progression of T2D, as it accounts for the majority (∼85%) of whole-body glucose disposal.^3^ Obesity is the primary cause of insulin resistance and is present in >90% of patients with T2D.^4,5^ As such, there is a critical need to identify the cellular and molecular factors mediating the progression of obesity-related skeletal muscle insulin resistance to overt T2D.

Mitochondria are dynamic organelles that maintain networking through processes of fission, fusion, biogenesis, and mitophagy.^6^ Maintenance of these networks is required to sustain mitochondrial function on multiple levels, including energy production and stress responsiveness 7 Mitochondrial Rho GTPase 1 (Miro1) is an evolutionarily conserved atypical GTPase residing on the outer mitochondrial membrane with calcium-binding motifs and a cytoplasmic facing c-terminal transmembrane domain.^8^ Early investigations of the Miro proteins revealed influential roles on mitochondrial morphology and movement in cells with compartmentalized transport requirements such as neurons.^9–11^ In contrast, the mitochondrial network configuration of muscle-specific MIro1 deficient flies was entirely intact despite a complete loss of climbing ability,^12^ indicative of previously unidentified roles in cell signaling and metabolism. We and others have shown that skeletal muscle lipid accumulation directly contributes to the onset and progression of insulin resistance by early and sustained activation of mitochondrial fission mediated by dynamin-related protein 1 (DRP1).^13–15^ DRP1 is critical to normal mitochondrial function, structure and turnover. Patients with T2D exhibit numerous mitochondrial defects, including decreased mitochondrial content and size, and altered cristae structure.^16–18^ Miro1 is required to anchor DRP1 to promote mitochondrial fission and restrict mitochondrial transport.^19^ However, the role and regulation of skeletal muscle Miro1 and its relationship to lipid-induced insulin resistance is entirely unknown.

Here, we examined the role and regulation of skeletal muscle Miro1 in patients and mice with obesity and T2D. We discovered that Miro1 accumulates in the skeletal muscle of mice and patients with obesity and hyperglycemia which is caused, in part, by defective mitochondrial protein kinase B (AKT) signaling and subsequently, inadequate AKT: Miro1 protein interaction. Improving skeletal muscle insulin sensitivity by means of exercise training relieved Miro1 accumulation while synergistically enhancing skeletal muscle oxidative capacity and restricting lipid accrual in patients with T2D. Congruently, skeletal muscle-specific deletion of Miro1 in mice or C2C12 cells enhances mitochondrial oxidative capacity, glucose transporter 4 (GLUT4) exocytosis, and glucose disposal. Taken together, these data support the role and regulation of Miro1 in glucose homeostasis and T2D.

## RESULTS

### Miro1 accumulation is associated with insulin resistance in patients and mice with obesity and type 2 diabetes

DRP1- and PINK1-mediated regulation of mitochondrial quality control has been shown to regulate Miro1 stability.^20^ In the context of Parkinson’s disease, defects in mitophagy lead to accumulation of Miro1 in skin fibroblasts.^21^ We have previously shown that activation of PINK1 contributes to lipid-induced insulin resistance in humans.^13^ Based upon these findings, we sought to examine skeletal muscle Miro1 expression across the glucoregulatory spectrum. To achieve this, we conducted a retrospective analysis of patients previously assigned to one of three groups based upon the following: healthy weight (HW: BMI 18-25 kg/m^2^ with no prior history of T2D or use of glucose-lowering medications), obesity without T2D (Ob: BMI 25-50 kg/m^2^ without T2D), or obesity with T2D (T2D: BMI 25-50 kg/m^2^ with T2D). Following screening, all patients underwent a three-day inpatient stay with a skeletal muscle biopsy followed by a single-stage, 180 min hyperinsulinemic-euglycemic clamp (**Fig. 1A**). Patients with T2D had elevated HbA1c, and impaired glucose disposal and insulin sensitivity compared to HW and Ob (**Fig. 1B-C**, **Table 1**). Furthermore, Miro1 protein accumulation was only observed in the skeletal muscle of patients with T2D (**Fig. 1D-E**), indicating a link between increased Miro1 expression and insulin resistance but not with obesity alone.

**Figure 1.**
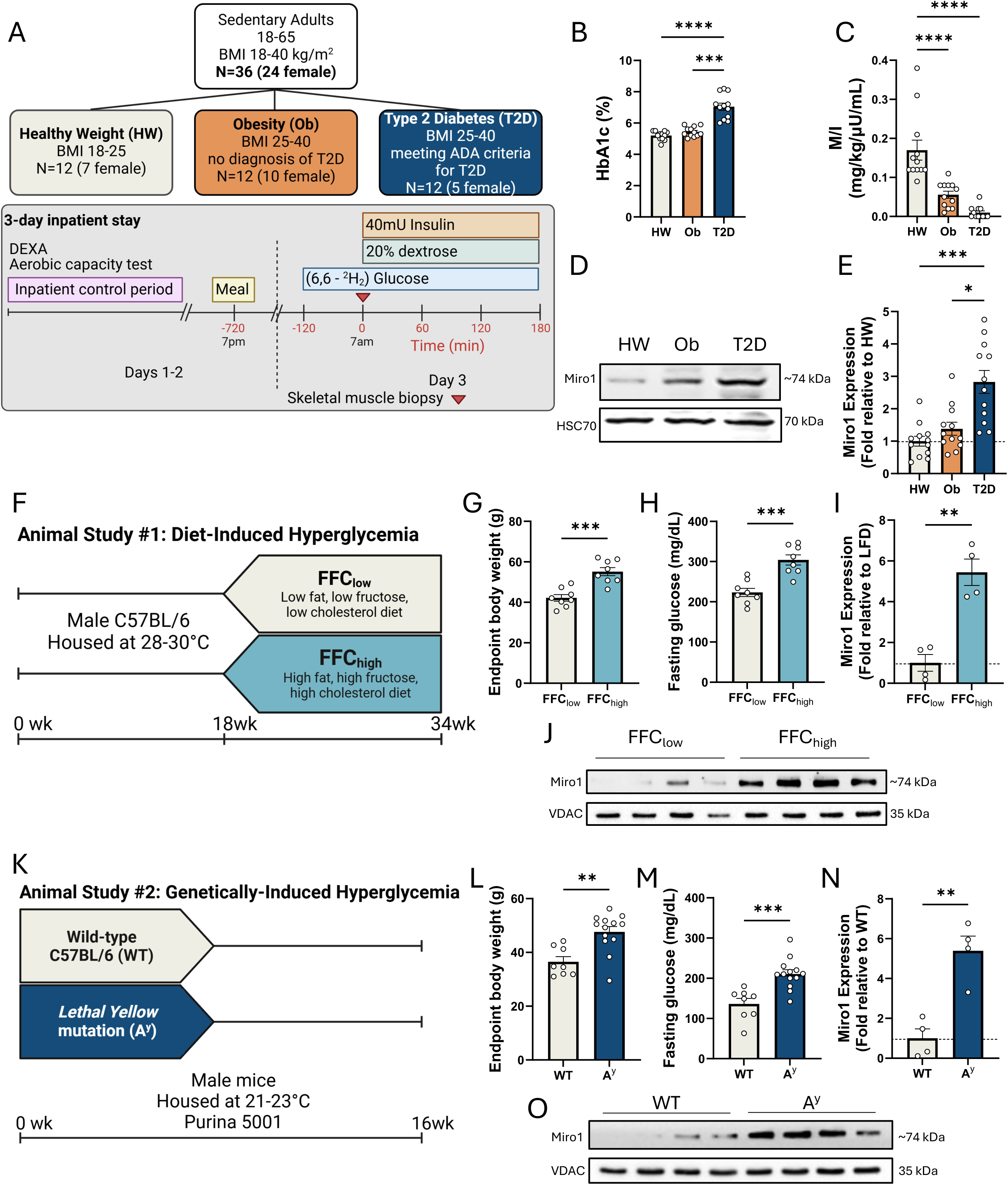
Miro1 accumulation is associated with insulin resistance in patients and mice with obesity and Type 2 Diabetes. (**A**) Overview of human cross-sectional study design. **(B-C**) Fasting HbA1c and glucose disposal rate relative to insulin during hyperinsulinemic-euglycemic clamps. (**D-E**) Western blot and quantification of fold change relative to HW for Miro1 expression in human skeletal muscle tissue from HW, Ob, and T2D patients (n=12/group). (**F**) Study design of diet-induced hyperglycemia in mice. (**G-H**) Endpoint body weight and fasting glucose of mice maintained on a low (FFClow) or high fat, fructose, and cholesterol diet (FFChigh) for 16 weeks (n=8/group). (**I-J**) Western blot and quantification of fold change relative to FFClow for Miro1 expression in skeletal muscle from mice fed a FFClow or FFChigh diet (n=4/group). (**K**) Study design of genetically induced hyperglycemia in mice. (L-M) Endpoint body weight and fasting glucose of wild-type C57BL/6 (WT) or A^y^ knockout (A^y^) mice (n=8/group). (**N-O**) Western blot and quantification of fold change relative to WT for Miro1 expression in skeletal muscle from WT and A^y^ mice (n=4/group). See also ***Figure S1***. Data are represented as mean ± SEM, *p < 0.05, **p < 0.01, ***p < 0.001, ****p < 0.0001, and were statistically compared by Krusal-Wallis test with Dunn’s multiple comparisons (B and E), one-way ANOVA (C) or Student’s t test (G-I and L-N).

**Table 1.** Cross-sectional cohort participant characteristics. Patient characteristics in individuals with Healthy Weight (HW), Obesity (Ob), or Type 2 Diabetes (T2D). Data are mean ± SD. BMI, body mass index; HDL, high-density lipoprotein; LDL, low-density lipoprotein.

| Characteristics |  |  |  | P value |  |  |
| --- | --- | --- | --- | --- | --- | --- |
|  | HW (N=12) | Ob (N=12) | T2D (N=12) | HW vs. Ob | HW vs. T2D | Ob vs. T2D |
| **Age (yrs)** | 28.0 ± 8.3 | 34.9 ± 7.5 | 50.8 ± 8.5 | 0.1066 | **<0.0001** | **<0.0001** |
| **Sex (M, %)** | 5 (41.7) | 2 (16.7) | 7 (58.3) | 0.4137 | 0.6711 | 0.097 |
| **Body Weight (kg)** | 66.4 ± 10.7 | 96.8 ± 14.4 | 104.3 ± 17.3 | **<0.0001** | **<0.0001** | 0.4261 |
| **BMI (kg/m<sup>2</sup>)** | 22.4 ± 1.7 | 31.6 ± 3.8 | 36.5 ± 6.5 | **<0.0001** | **<0.0001** | **0.0297** |
| **Triglyceride (mg/dL)** | 94.6 ± 89.4 | 132.6 ± 83.2 | 142.2 ± 73.7 | 0.5026 | 0.3448 | 0.9563 |
| **Total Cholesterol (mg/dL)** | 178.1 ± 42.4 | 184.7 ± 35.1 | 178.1 ± 39.8 | 0.9112 | >0.9999 | 0.9112 |
| **HDL Cholesterol (mg/dL)** | 62.7 ± 16.2 | 43.9 ± 8.2 | 45.5 ± 10.4 | **0.0016** | **0.0039** | 0.944 |
| **LDL Cholesterol (mg/dL)** | 96.7 ± 40.6 | 114.2 ± 19.0 | 104.1 ± 30.7 | 0.3687 | 0.8308 | 0.7137 |
| **VO<sub>2</sub>PEAK (mL/kg/min)** | 40.0 ± 10.4 | 30.5 ± 7.2 | 20.2 ± 3.6 | **0.0116** | **<0.0001** | **0.0062** |
| **Fasting Glucose (mg/dL)** | 87.6 ± 5.6 | 86.5 ± 7.4 | 167.4 ± 51.9 | 0.9956 | **<0.0001** | **<0.0001** |
| **Fasting Insulin (μU/mL)** | 6.7 ± 2.4 | 14.0 ± 8.3 | 17.6 ± 8.8 | **0.0444** | **0.0018** | 0.4209 |

To confirm these findings, we evaluated skeletal muscle Miro1 expression in various mouse models of T2D. First, we compared mice on a high fat, high fructose, and high cholesterol (FFChigh) diet (**Fig. 1F**), which caused obesity, hyperglycemia, and subsequently, skeletal muscle Miro1 accumulation compared to mice on a low fat, low fructose, low cholesterol (FFClow) diet (**Fig. 1G-J**). To determine if Miro1 accumulation in mice with obesity and hyperglycemia was explained by diet alone, we investigated Miro1 expression in two monogenic mouse models of T2D compared to littermate controls. We observed that A^y^ and Lepr^db-/-^ mice maintained on a low-fat diet exhibit similar accumulation of skeletal muscle Miro1 compared to FFChigh mice (**Fig. 1K-O, Supplementary** Figure 1A-C). Taken together, these data support skeletal muscle Miro1 accumulation in humans and mice with obesity-related T2D.

### Defective mitochondrial AKT translocation mediates Miro1

We next sought to determine the etiology of skeletal muscle Miro1 accumulation in the context of hyperglycemia. Given our observations that Miro1 selectively accumulates in the background of T2D, we hypothesized that altered skeletal muscle glucoregulatory function may cause Miro1 accumulation. Since insulin is the primary endocrine mediator of intracellular glucose disposal in skeletal muscle, we first evaluated the extent to which exogenous exposure may affect Miro1 expression. Acute exposure of C2C12 cells to insulin decreased Miro1 expression, indicating a potential link between insulin receptor signaling and regulation of Miro1 status on the outer mitochondrial membrane (**Fig. 2A- B**). In the canonical insulin signaling cascade, AKT phosphorylation in the cytosol leads to glucose uptake by facilitating the translocation of GLUT4 to the plasma membrane.^22^ However, it has been previously shown that AKT phosphorylation at the insulin-dependent threonine 308 residue rapidly accumulates in mitochondria.^23^ We evaluated these observations by measuring AKT^T308^ phosphorylation from isolated skeletal muscle mitochondria following in vivo infusion of exogenous insulin. T2D patients exhibited a near-complete absence of mitochondrial AKT^T308^ phosphorylation when compared to patients with obesity alone (**Fig. 2C-D**). Given that Miro1 accumulation in the mitochondria is associated with skeletal muscle insulin resistance and that defective AKT translocation results in mitochondrial AKT accumulation in T2D, we hypothesized that AKT confinement to the mitochondria in T2D is related to Miro1 accumulation. To test this, we performed immunofluorescence analysis of C2C12 cells, where co-localization of AKT to Miro1 in TOM20-expressing regions was relatively low under basal conditions, which increased to ∼60% in the presence of insulin (**Fig. 2E-G)**. To further support this, we performed co-immunoprecipitation with mitochondrial AKT and Miro1 in mouse gastrocnemius to determine if the proteins interact directly and whether this is altered by the presence of insulin. We observed that under basal conditions, small but measurable amounts of Miro1 were bound to AKT (**Fig. 2H-J**), presumably explained by the lesser quantity of mitochondrial AKT observed in the absence of insulin. In the presence of insulin, there was a significant increase in the protein interaction between Miro1 and mitochondrial AKT, largely attributable to the increased AKT expression in the presence of insulin. We next sought to determine if mitochondrial AKT translocation was required to restrict Miro1 accumulation. To address this, C2C12 cells were exposed to MK-2206, a highly selective pan Akt inhibitor,^24^ in the presence or absence of insulin. We observed that in the absence of insulin, MK-2206 did not alter Miro1 expression, whereas in the presence of insulin, inhibition of AKT by MK-2206 caused rapid Miro1 accumulation (**Fig. 2K-M**). Taken together, these data indicate that Miro1 accumulation in T2D is mediated by defective mitochondrial AKT translocation in response to insulin.

**Figure 2.**
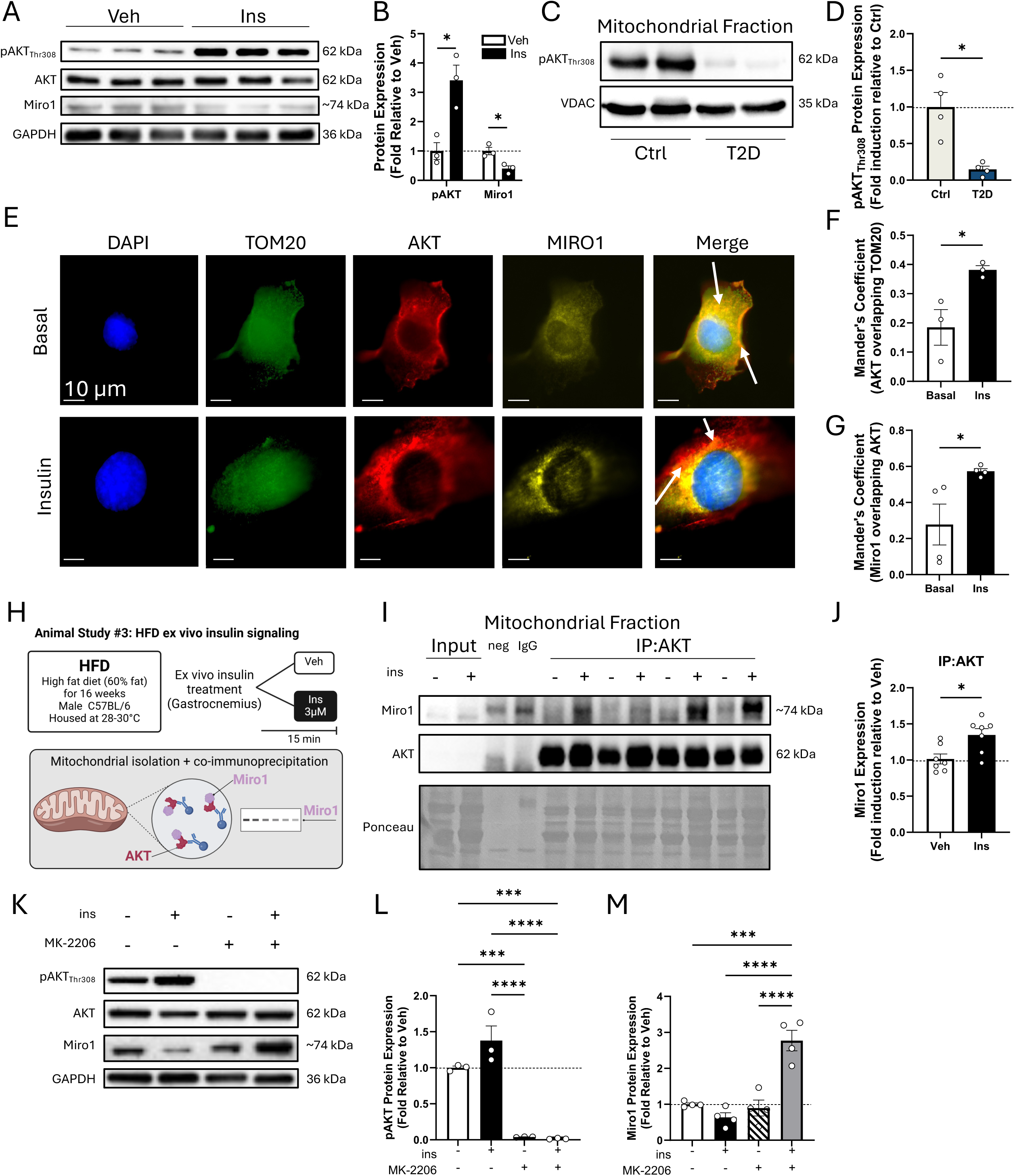
Defective mitochondrial AKT translocation mediates Miro1 accumulation. (**A-B**) Western blot and quantification of fold change relative to vehicle for AKT and Miro1 from C2C12 myoblasts with and without 1mM insulin treatment (n=3 biological replicates/condition). (**C-D**) Western blot and quantification of fold change relative to healthy control patients for mitochondrial AKT from human skeletal muscle mitochondria (n=4/group). (**E**) Immunofluorescence staining of AKT (red), Miro1 (yellow), DAPI (blue), TOM20 (green), and the resulting merged stack in C2C12 myoblasts treated with and without insulin. Scale = 10μm. (**F**) Quantification of Mander’s coefficient for co-localization of AKT to TOM20 (n=3 biological replicates/condition) and (**G**) Miro1 to AKT in TOM20 expressing regions in the presence or absence of insulin (n=4 biological replicates/condition). (**H**) Study design of coimmunoprecipitation in ex-vivo insulin treated gastrocnemius muscle mitochondria from mice fed a high fat diet (HFD). (**I-J**) Western blot and quantification of fold change relative to vehicle for gastrocnemius mitochondrial Miro1 co-expressed with AKT with and without ex-vivo insulin treatment in mice fed a HFD (n=8 biological replicates/condition). (**K-M**) Western blot and quantification of fold change relative to non-treatment control for AKT and Miro1 in C2C12 myoblasts with and without insulin and MK-2206 treatment (n=3 biological replicates/condition). Data are represented as mean ± SEM, *p < 0.05, **p < 0.01, ***p < 0.001, ****p < 0.0001, and were statistically compared by Student’s t test (B, D, F, and J), Mann-Whitney test (G), or one- way ANOVA (L and M). For panel D, the p-value was adjusted for unequal variance by Welch’s correction.

### Exercise training relieves skeletal muscle mitochondrial hyper-fragmentation and Miro1 aggregation in patients with obesity and Type 2 Diabetes

Given our observations that Miro1 selectively accumulates in the skeletal muscle of patients with T2D, which is caused, at least in part, by insufficient interaction between Miro1 and the insulin signaling pathway, we hypothesized that treatments that improve peripheral insulin sensitivity may relieve Miro1 accumulation. Exercise training is a first-line lifestyle therapy for managing T2D^25^ and is well known to resensitize skeletal muscle glucoregulatory and mitochondrial function.^26^ To address this, we conducted a parallel arm, open-label, randomized controlled trial comparing exercise training to standard of care in patients with established obesity and T2D (**Fig. 3A, Fig. S2**). Patients randomized to standard of care (Ctrl) continued with routine medical management for T2D for 12 weeks, while also receiving counseling on lifestyle management of disease. Patients randomized to exercise training (Ex) underwent a 12-week aerobic training program consisting of moderate-intensity, steady state treadmill walking/jogging for 5 days per week, 60 minutes per session. All patients underwent an inpatient stay consisting of a two-step hyperinsulinemic-euglycemic clamp and skeletal muscle biopsies before and after the intervention period (**Fig. 3A**). Age, sex, body weight, diabetes duration, number of glucose-lowering medications, and key demographic variables were comparable at baseline (**Table 2**). No serious adverse events were reported. Ex increased VO2peak and lean mass while reducing fat mass relative to Ctrl (**Table 3**). At the muscular level, ultrastructural analyses of the contractile apparatus revealed that exercise increased myofibrillar connectivity as evidenced by the greater number of sarcomere branches after 12 weeks (**Fig. 3B-C**). Additionally, the lipid droplet volume per cell significantly decreased following exercise training (**Fig. 3D-E, Supplementary** Fig. 3A-F). Peripheral insulin sensitivity was improved in Ex but not Ctrl following the intervention (**Fig. 3F**), as well as GLUT4 translocation to skeletal muscle membrane (**Fig. 3G-H)**. Ex lowered Miro1 expression which remained unchanged in Ctrl (**Fig. 3G,I**). Ex reduced skeletal muscle DRP1 phosphorylation (**Fig. 3G,I**), as well as mediators responsible for recruiting DRP1 to the mitochondrial surface, such as mitochondrial outer membrane protein 1 (Mid51) and 2 (Mid49) (**Fig. 3J-K**). Since DRP1 is required for mitochondrial fission and we have previously observed improvements in mitochondrial elongation in response to exercise training,^26^ we assessed mitochondrial ultrastructure and morphology by transmission electron microscopy. At baseline, Ex and Ctrl skeletal muscle mitochondria were relatively small and punctate, with consistent distribution across the subsarcolemmal and intermyofibrillar regions (**Fig. 3L**). Following the intervention period, mitochondria remained largely punctate and fragmented in Ctrl, whereas Ex improved the sphericity and elongation of individual and networked mitochondria, resulting in a significant improvement in the fragmentation index (**Fig. 3M**). Based on our findings that exercise training improved mitochondrial dynamics, we explored changes in other protein mediators of skeletal muscle biogenesis and quality control. We did not observe increases in PGC1α, MFN1, MFN2, OPA1, PINK1, or Parkin expression in Ex compared Ctrl following intervention (**Supplementary** Fig. 4A-B). We observed decreased expression of LC3, which acts as a signaling protein to remove damaged mitochondria,^27^ in patients following Ex compared to Ctrl (**Fig. 3J-K**).

**Figure 3.**
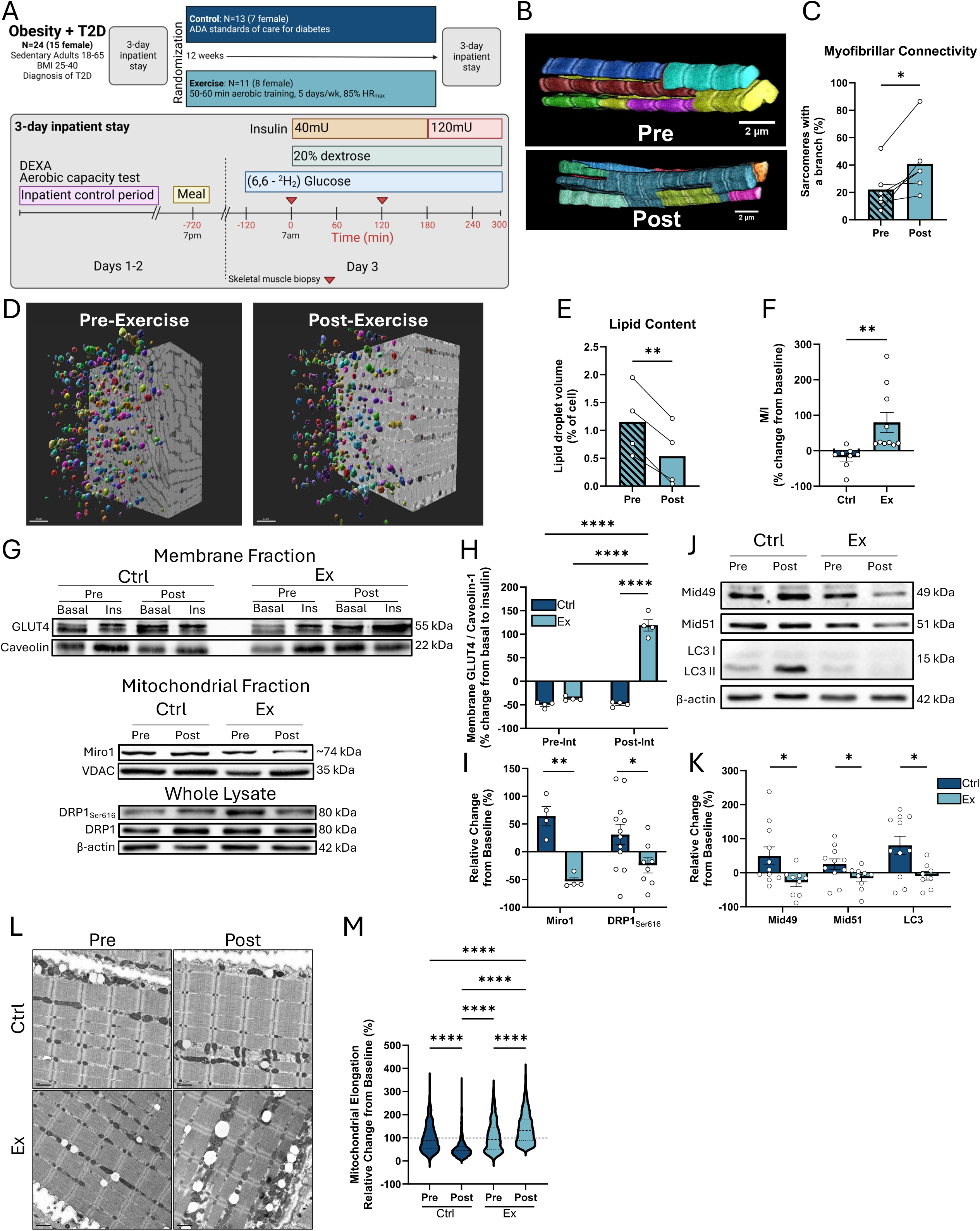
Exercise training relieves skeletal muscle mitochondrial hyper-fragmentation and Miro1 aggregation in patients with obesity and Type 2 Diabetes. (**A**) Overview of randomized controlled trial study design. See also ***Figure S2***. (**B-C**) Ultrastructural analyses of skeletal muscle contractile apparatus and quantification of myofibrillar connectivity based on percent of sarcomeres with a branch before and after exercise training (n=5). Different colors are representative of individual sarcomeres. Scale bars represent 2μm. (**D-E**) Ultrastructure analyses of skeletal muscle lipid droplets and quantification of lipid droplet content before and after exercise training (n=4). Different colors are representative of individual lipid droplets. Scale bars represent 300μm. See also ***Figure S3.*** (**F**) Relative change from baseline in insulin sensitivity during hyperinsulinemic-euglycemic clamp in Ctrl and Ex groups. (**G-I**) Western blots and quantification from human skeletal muscle samples in Ctrl and Ex groups before and after 12-week intervention. Relative changes in GLUT4 expression in response to insulin relative to basal condition from skeletal muscle membrane fractionation before and after Ctrl or Ex (n=4/group). Relative changes from baseline in mitochondrial Miro1 (n=4/group) and whole lysate DRP1 (n=8-12/group) expression in human skeletal muscle samples before and after 12-week intervention. (**J-K**) Western blots and quantification from human skeletal muscle samples in Ctrl and Ex groups before and after 12-week intervention. Relative changes from baseline in Mid49, Mid51, and LC3 expression before and after 12-week intervention (n=8-12/group). See also ***Figure S4.*** (**L-M**) Electron microscopy images of human skeletal muscle samples from Ctrl and Ex patients before and after 12- week intervention (n=4/timepoint/group) and mitochondrial fragmentation index quantified from all distinguishable mitochondria per field. Data are represented as mean and individual change (C and E), mean ± SEM (F, H, I, and K), or mean and quartiles (M), *p < 0.05, **p < 0.01, ****p < 0.0001, and were statistically compared by Student’s t test (C, E, F, and I), two-way repeated measures ANOVA with Holm-Šídák’s multiple comparisons test (H), or Kruskal-Wallis ANOVA with Dunn’s multiple comparison’s test (M), or Welch’s t-test (K). For panel F, the p-value was adjusted for unequal variance by Welch’s correction.

**Table 2.**
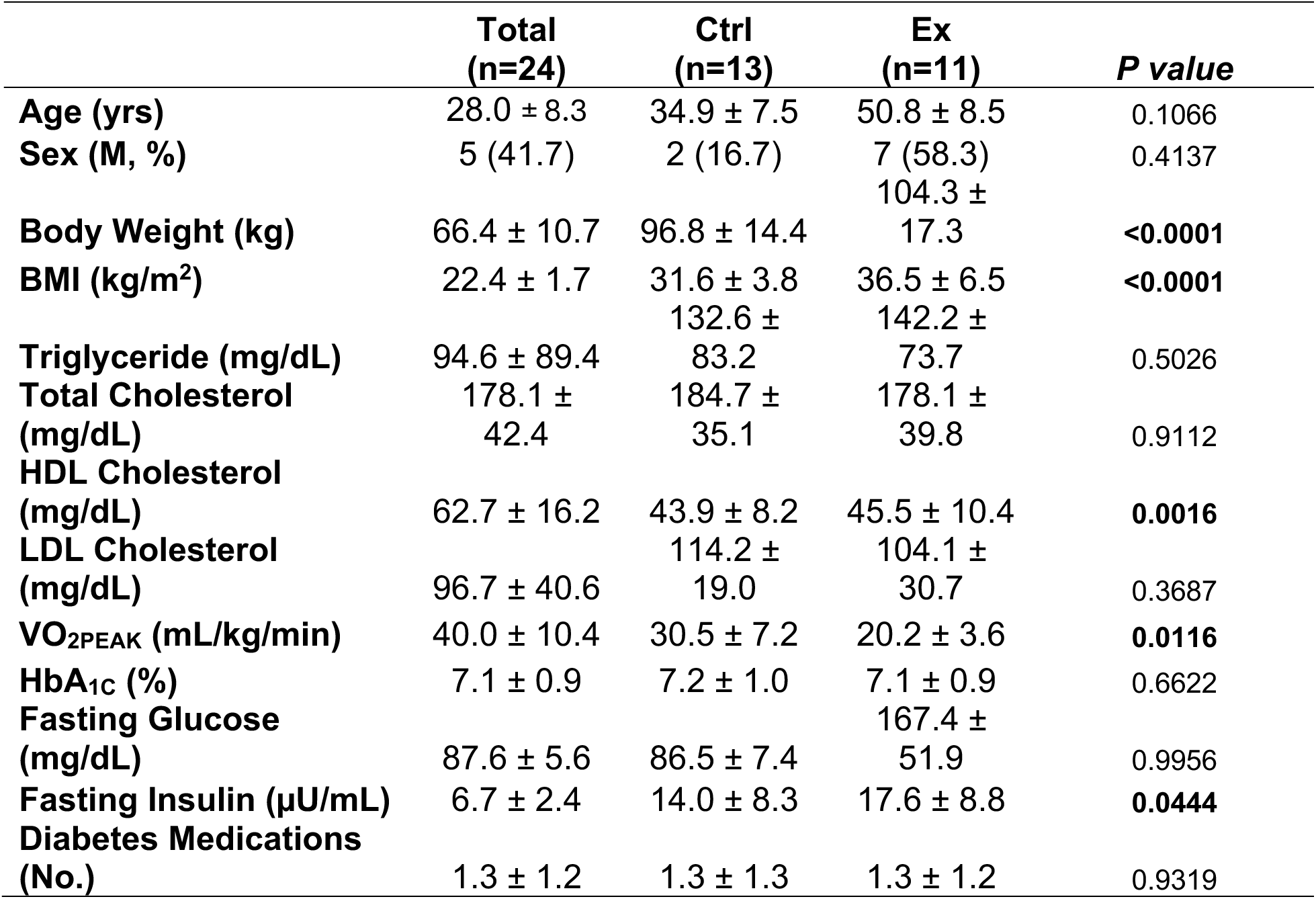
Baseline characteristics of patients with obesity and Type 2 Diabetes in Exercise RCT. Patient characteristics at baseline in control (Ctrl) and exercise (Ex) groups. Data are mean ± SD. BMI, body mass index; HDL, high-density lipoprotein; LDL, low-density lipoprotein.

**Table 3.** Changes from baseline in control (Ctrl) and exercise (Ex) patients. Data are shown as relative change from baseline (post-pre) ± standard error. *P* value is (post-pre Ex) vs (post-pre Ctrl). BMI, body mass index.

|  | Ctrl | Ex | P value |
| --- | --- | --- | --- |
| **Body Weight (kg)** | -0.7773 ± 1.1015 | -3.2574 ± 0.9714 | 0.1060 |
| **BMI (kg/m<sup>2</sup>)** | 0.09822 ± 0.4885 | -1.0308 ± 0.4412 | 0.1004 |
| **Fat Mass (kg)** | 0.1524 ± 0.3551 | -1.2872 ± 0.3179 | **0.0063** |
| **Lean Mass (kg)** | -0.4109 ± 0.444 | 0.03861 ± 0.4021 | 0.4610 |
| **VO<sub>2PEAK</sub> (mL/kg/min)** | -0.267 ± 0.6683 | 2.2807 ± 0.5599 | **0.0081** |
| **Fasting Glucose (mg/dL)** | -13.275 ± 32.321 | -12.45 ± 29.112 | 0.9552 |
| **Fasting Insulin (μU/mL)** | 0.729 ± 4.438 | -3.837 ± 5.979 | **0.0389** |

In line with differences in skeletal muscle mitochondrial structure and networking following exercise, we then investigated whether these improvements were linked to enhancements in mitochondrial function. Using high resolution respirometry to assess maximal oxidative phosphorylation (OXPHOS) and electron transfer (ET) capacity in isolated mitochondria from skeletal muscle of patients with T2D (**Fig. 4A**), we observed increases in maximal NADH (N)-, succinate (S)-, palmitoyl carnitine (F)-, and complex III (CIII)-linked OXPHOS and complex IV (CIV)-linked ET in Ex compared to Ctrl (**Fig. 4B**). Acceptor control ratio was also increased following Ex (**Fig. 4C**), collectively indicating increases in maximal skeletal muscle OXPHOS and improved coupling of substrate oxidation to ATP phosphorylation in Ex compared to Ctrl. N-linked ET was also increased following Ex, despite similarities in S- and CIII-linked ET capacity between groups (**Supplementary** Fig. 5). Based on the observed improvements in OXPHOS capacity following Ex, we wanted to determine if the changes were consistent with physiological improvements in respiratory control. To accomplish this, we employed the creatine kinase clamp,^28^ which enables evaluation of the conductivity of the respiratory system over a wide range of free energy states through external control of the extramitochondrial ATP:ADP ratios (**Fig. 4D**).^28^ Consistent with our maximal OXPHOS assessments, Ex increased N- and S-linked conductance and Vmax compared to Ctrl (**Fig. 4E-F**). Taken together, these data suggest that exercise training relieves skeletal muscle hyper-fragmentation and Miro1 accumulation while enhancing respiratory capacity and conductance in patients with T2D.

**Figure 4.**
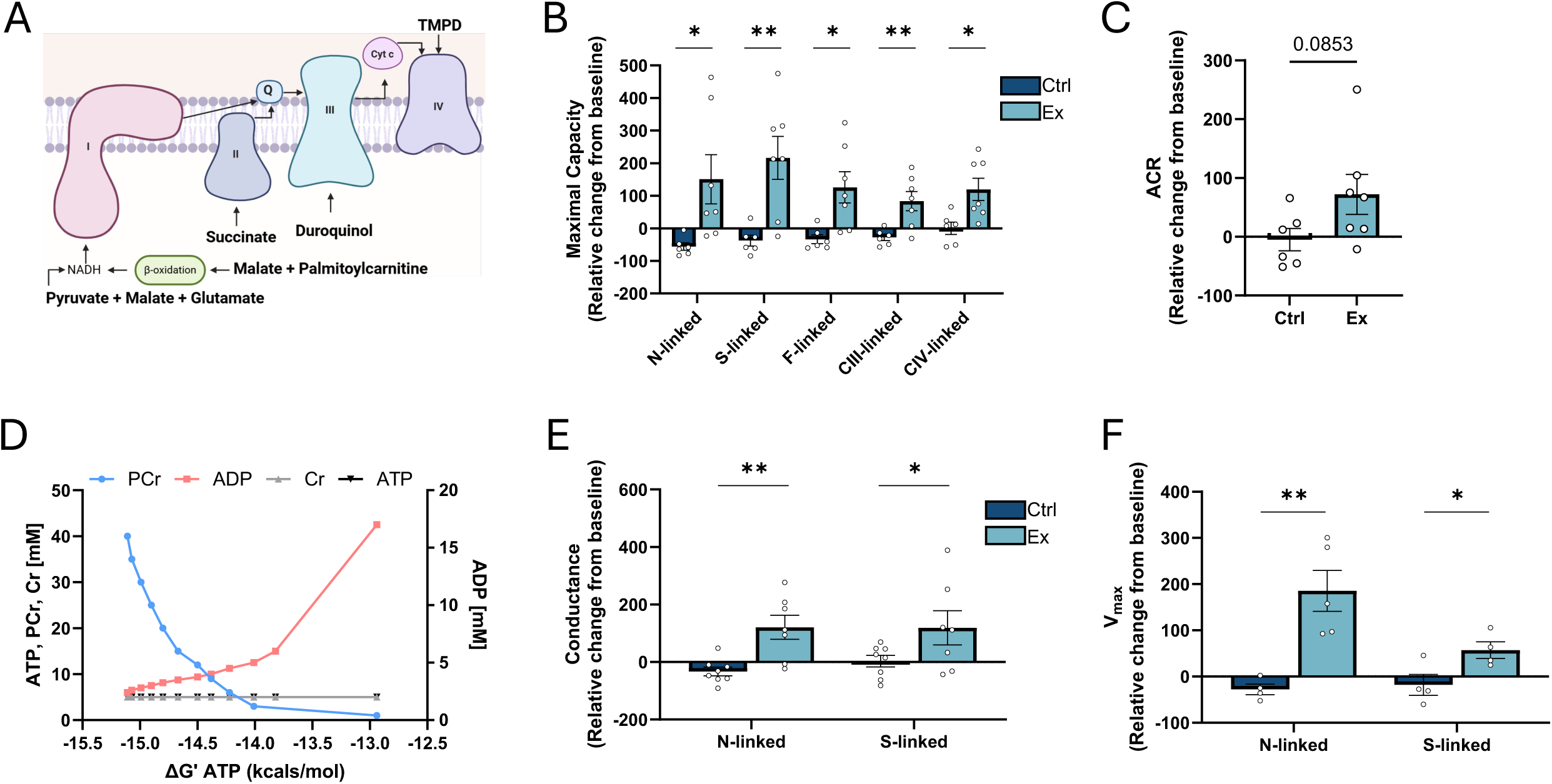
Exercise training enhances skeletal muscle mitochondrial capacity and conductance in patients with obesity and Type 2 Diabetes. (**A**) Overview of the experimental approach to evaluate mitochondrial respiratory capacity. (**B-C**) Relative changes from baseline in maximal NADH-linked (N-linked), succinate-linked (S-linked), fatty acid-linked (F-linked), complex III-linked (CIII-linked), and complex IV-linked (CIV-linked) mitochondrial oxidation and acceptor-control ratio (ACR) in Ctrl and Ex patients (n=6-7/group). See also ***Figure S5.*** (**D**) Representative plot illustrating the change in free energy of ATP production (ΔG’ ATP) in response to phosphocreatine (PCr) titration using the bioenergetic creatine kinase clamp technique. (**E-F**) Relative changes from baseline in N-linked and S-linked mitochondrial conductance and Vmax (n=4- 7/group). Data are represented as mean ± SEM. *p < 0.05, **p < 0.01 by Student’s t test.

### Miro1 negatively regulates skeletal muscle insulin action and oxidative function in mice with obesity and hyperglycemia

Since exercise promotes multi-organ, pleotropic effects on physiological function, we sought to mechanistically tease apart the contribution of skeletal muscle Miro1 to maintaining glucoregulatory function in vivo. To achieve this, we generated a skeletal-muscle specific Miro1-deficient mouse model (**Supplementary** Fig. 6A) by crossing homozygous Miro1-floxed mice with the ACTA1-Cre mice, resulting in spatially controlled Cre-mediated deletion of Miro1 (Miro1^SkM-/-^). Miro1^SkM-/-^ mice exhibited ∼70% reduction in Miro1 protein and >90% reduction in gene expression, which was restricted to skeletal muscle tissues (**Fig. 5A-B**, **Supplementary** Fig. 6B). Miro1^SkM-/-^ mice were fertile, viable, and did not display overt development defects or deficiencies (**Supplementary** Fig. 6C). To determine effects on insulin sensitivity, Miro1^SkM-/-^ and Miro1^fl/fl^ were placed on a high-fat diet (HFD) for 16 weeks, followed by evaluation of peripheral glucose disposal by intraperitoneal insulin tolerance testing and harvesting of muscle to evaluate insulin signaling (**Fig. 5C**). Body weight and food intake were similar in Miro1^SkM-/-^ mice relative to littermate controls (**Fig. 5D, Supplementary** Fig. 7A-C), while glucose disposal in response to insulin was improved in Miro1^SkM-/-^ mice (**Fig. 5E-F**). Fasting glucose, fasting insulin, and HOMA-IR were also lower in Miro1^SkM-/-^ mice (**Fig. 5G-I**). To determine the skeletal muscle-specific effects of Miro1 deletion on insulin resistance, gastrocnemius muscles were exposed to rapid acting insulin ex vivo and tissues were lysed to assess insulin signaling. Miro1^SkM-/^- mice had increased AKT and AS160 signaling in skeletal muscle compared to Miro1^fl/fl^ mice (**Fig. 5J-L**). Membrane-bound GLUT4 in response to insulin was also higher in Miro1^SkM-/-^ (**Fig. 5J, M**). OXPHOS capacity and conductance were also assessed in the quadricep muscle from Miro1^SkM-/^- and Miro1^fl/fl^ mice to evaluate the direct effects of respiratory function. Miro1^SkM-/-^ mice exhibited increased N- and S-linked OXPHOS (**Fig. 5N**), as well as increased N- and S-linked maximal respiration and conductance (**Fig. 5O-P**).

**Figure 5.**
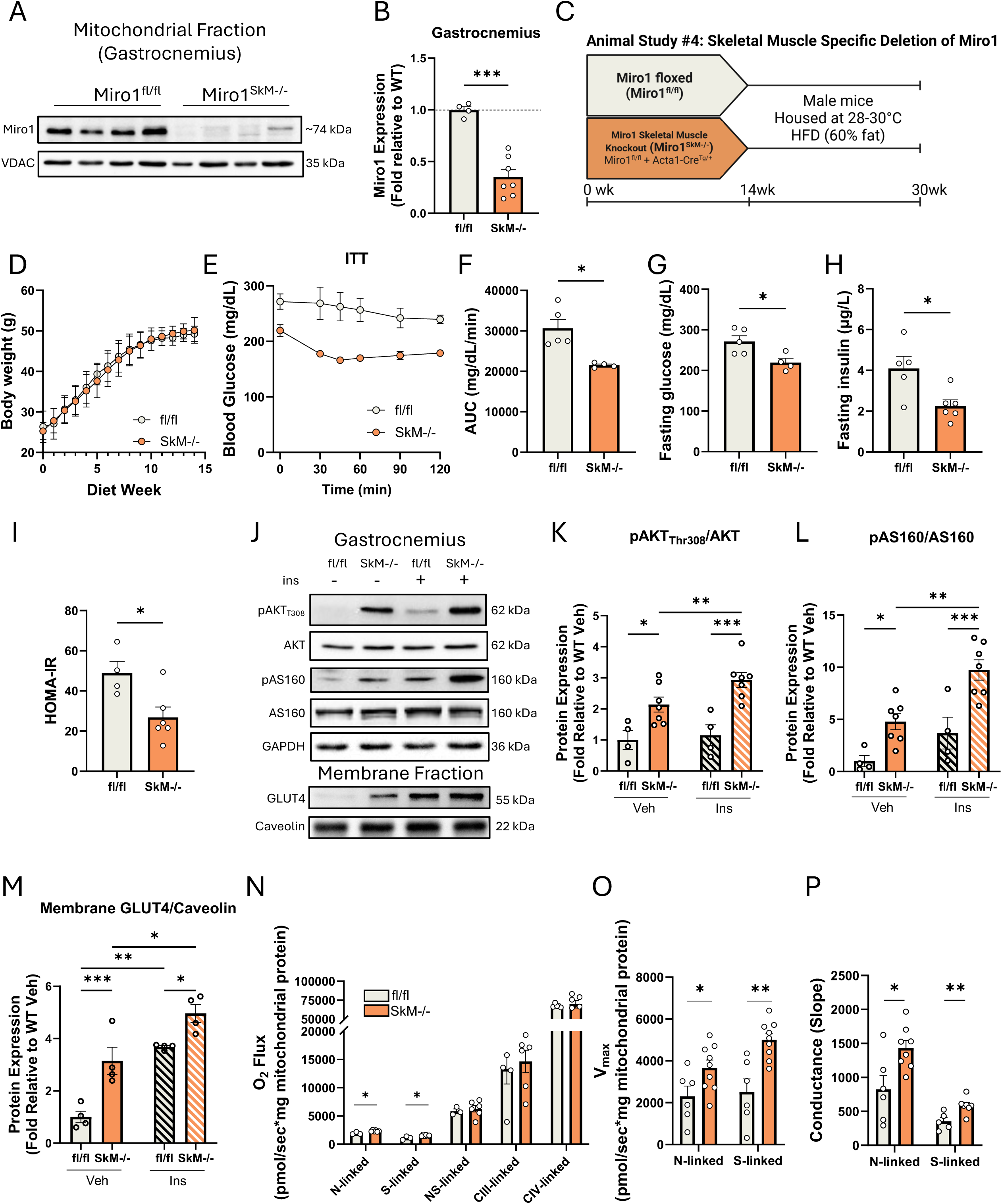
Miro1 negatively regulates skeletal muscle insulin action and oxidative capacity in mice with obesity and hyperglycemia. (**A-B**) Western blot and quantification of skeletal muscle Miro1 expression in Miro1 floxed (fl/fl) and skeletal muscle Miro1 knockout (SkM-/-) mice (n=4-7/group). See also ***Figure S6.*** (**C**) Study design of Miro1 fl/fl and SkM-/- mice on a high-fat diet (HFD). (**D**) Body weight curves of Miro1 fl/fl and SkM-/- mice from weekly measurements during experimental diet (n=8-10/group). See also ***Figure S7.*** (**E-F**) Responses to an intraperitoneal insulin tolerance test and resulting area under the curve (AUC) in Miro1 fl/fl and SkM-/- mice (n=4-5/group). (**G-I**) Fasting glucose, insulin, and calculated HOMA- IR in Miro1 fl/fl and SkM-/- mice (n=4-5/group). (**J-M**) Western blot and quantification of fold change relative to fl/fl vehicle for gastrocnemius with and without ex-vivo insulin treatment in Miro1 fl/fl and SkM-/- mice (n=4-7/group) for insulin signaling proteins AKT, AS160, and membrane-bound GLUT4. (**N**) Maximal NADH-linked (N-linked), succinate- linked (S-linked), convergent NADH- and succinate-linked (NS-linked), complex III-linked (CIII-linked), and complex IV-linked (CIV-linked) mitochondrial oxidation in Miro1 fl/fl and SkM-/- mice (n=4-6/group). (**O-P**) N-linked and S-linked mitochondrial Vmax and conductance measured using creatine kinase clamps in Miro1 fl/fl and SkM-/- mice (n=4- 6/group). Data are represented as mean ± SEM, *p < 0.05, **p < 0.01, ***p < 0.001, and were statistically compared by Student’s t test (B, F, G, H, I, N, O, and P) or two-way repeated measures ANOVA with Holm-Šídák’s multiple comparisons test (D, K, L, and M). For panel F, the p-value was adjusted for unequal variance by Welch’s correction.

Given the possibility for paracrine or endocrine adaptations to Miro1 loss-of-function in mice, we validated these findings in vitro by inserting a Miro1-targeting (shMiro1) or empty vector (shEV) lentiviral construct into C2C12 skeletal muscle cells. Using this approach, we achieved loss of gene and protein expression in shMiro1 cells comparable to our animal model (**Fig. 6A-C, Supplementary** Fig. 8A), with no overt complications related to proliferation or cell structure (**Supplementary** Fig. 8B). To directly assess the effects on glucose uptake, cells were labeled with tritiated glucose in the presence or absence of insulin. shMiro1 cells exhibited a marked increase in glucose uptake in the presence and absence of insulin relative to shEV cells (**Fig. 6D**). GLUT4 translocation to the cell membrane in response to insulin was enhanced in shMiro1 cells (**Fig. 6E-F**). Congruent with our animal studies shMiro1 cells exhibited increased N- OXPHOS- OXPHOS relative to shEV compared to controls (**Fig. 6G**). Taken together, these data indicate that Miro1 negatively regulates skeletal muscle glucose uptake and bioenergetic function in vivo and in vitro.

**Figure 6.**
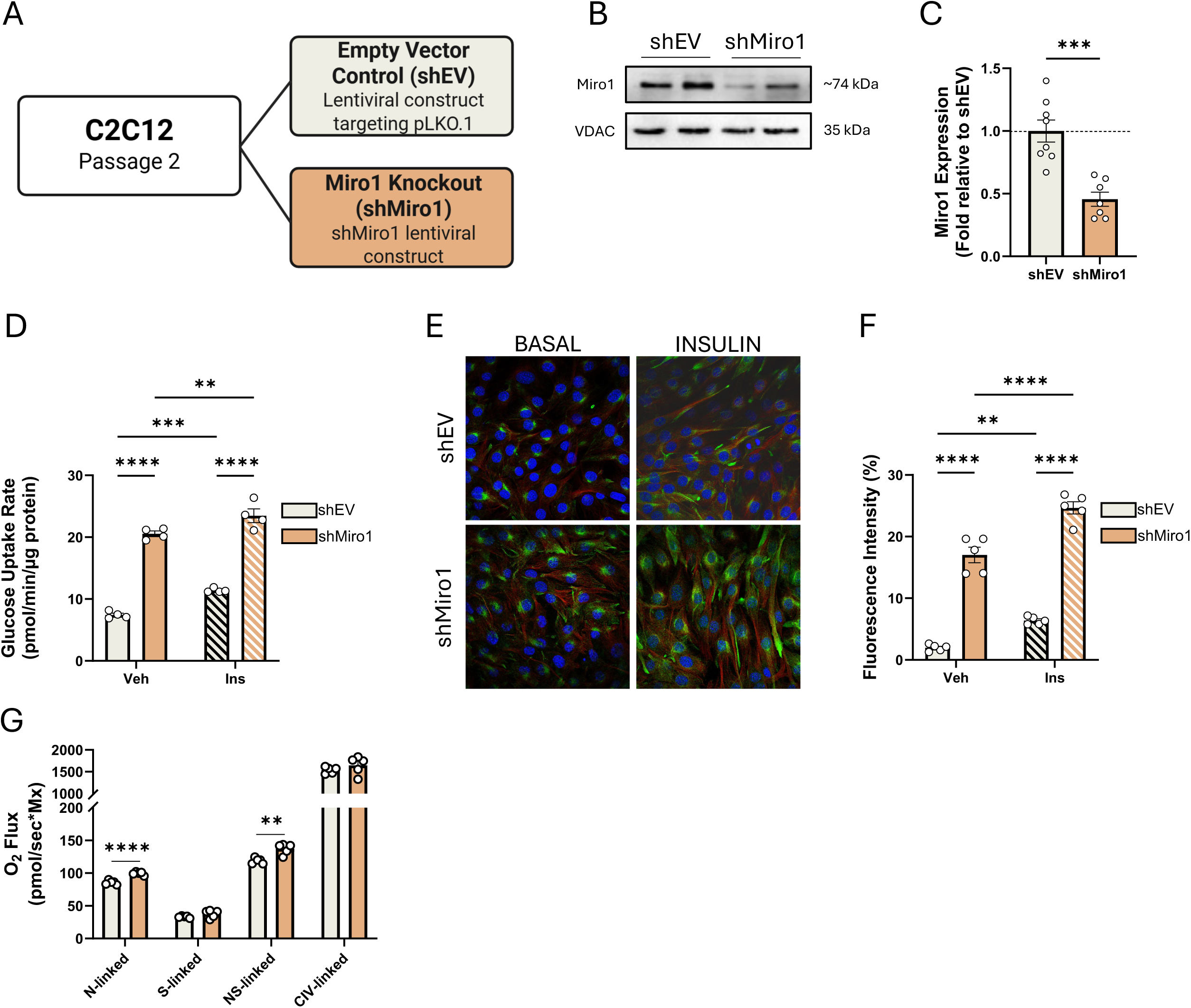
Miro1 negatively regulates intracellular glucose uptake and oxidative capacity in skeletal muscle cells. (**A**) Overview of design for lentiviral empty vector control (shEV) and Miro1 silencing (shMiro1) in C2C12 myoblasts. (**B-C**) Western blot and fold change quantification relative to shEV of Miro1 protein expression in shEV and shMiro1 C2C12 myoblasts (n=7-8 biological replicates/group). See also ***Figure S8.*** (**D**) [3-^3^H]glucose uptake in shEV and shMiro1 C2C12 cells treated with or without insulin (n=4 biological replicates/group/treatment). (**E-F**) Immunofluorescence staining for DAPI (blue), α-tubulin (red), and GLUT4 (green) and quantification of fluorescence intensity in shEV and shMiro1 cells treated with and without insulin (n=5 biological replicates/group/treatment). (**G**) Maximal NADH-linked (N-linked), succinate-linked (S-linked), convergent NADH- and succinate-linked (NS-linked), and complex IV-linked (CIV-linked) mitochondrial oxidation in shEV and shMiro1 C2C12 myoblasts (n=5 biological replicates/group/treatment). Data are represented as mean ± SEM, **p < 0.01, ***p < 0.001, ****p < 0.0001, and were statistically compared by Student’s t test (C and G) or two-way repeated measures ANOVA with Holm-Šídák’s multiple comparisons test (D and F).

## DISCUSSION

In this study, we investigated the role of Miro1 in skeletal muscle insulin resistance and glucose homeostasis in type 2 diabetes (T2D). Our findings reveal that Miro1 accumulates in the skeletal muscle of human patients and mouse models with obesity and hyperglycemia, driven in part by defective mitochondrial AKT signaling and impaired AKT-Miro1 interaction. Notably, exercise training alleviated Miro1 accumulation, improved mitochondrial oxidative capacity, and enhanced insulin sensitivity in individuals with T2D. Furthermore, skeletal muscle-specific deletion of Miro1 in mice and C2C12 cells enhanced mitochondrial function, insulin signaling, and glucose uptake, suggesting that Miro1 serves as a negative regulator of skeletal muscle insulin action. These findings support a previously unknown mechanism linking mitochondrial dynamics to insulin resistance and suggest that targeting Miro1 may offer therapeutic potential to enhance insulin sensitivity in T2D.

One of the principal observations of this study was the accumulation of skeletal muscle Miro1 in the context of obesity and T2D. Miro1 belongs to the Rho GTPase family, which encompasses a broad class of proteins that act as molecular switches by cycling bound GTP between the active and inactive states.^29^ Miro1 structure, function, and localization is atypical to the Rho GTPase family due to its calcium binding motifs as well as its mitochondrial outer membrane anchored GTPase domain in the C terminus region of the protein.^8,30^ Early functional studies of the protein revealed direct effects on mitochondrial dynamics, trafficking, calcium sensing, and inter-organelle connectivity.^20,31,32^ To this end, it was observed that loss of Miro1 function in neurons and astrocytes increased the rate of mitochondrial trafficking.^32,33^ In the context of Parkinson’s disease, mutations in PINK1 or Parkin restrict Miro1 degradation, leading to its accumulation, ultimately restricting axonal retrograde signaling and motility.^20^ However, unlike neuronal, skeletal muscle mitochondria reside in highly structured, reticulated networks without the persistent need for transport along axons.^34^ In drosophila, which expressed a highly conserved orthologous protein, skeletal muscle loss of Miro1 function renders the fly flightless without altering the mitochondrial network configuration,^12^ indicative of a cell autonomous effect with discrete regulatory properties. Similar to what is observed in Parkinson’s disease, Miro1 selectively accumulates in patients and mice with overt hyperglycemia, which was not explained by obesity alone, leading to the hypothesis that in skeletal muscle, this phenomenon may be glucose dependent.

AKT is a central mediator of insulin signaling, primarily known for facilitating glucose uptake through GLUT4 translocation via phosphorylation of key substrates such as AS160.^35^ However, emerging evidence suggests that AKT translocates to mitochondria in response to insulin^36^ which influences mitochondrial function and dynamics. Miro1, an outer mitochondrial membrane GTPase, has been implicated in mitochondrial trafficking and fission, interacting with key mitochondrial quality control proteins such as DRP1 and PINK1. Given that AKT phosphorylation at the mitochondria has been linked to metabolic control, it is plausible that its interaction with Miro1 modulates mitochondrial dynamics in response to insulin, potentially by altering fission-fusion balance or mitochondrial trafficking to meet cellular energy demands. The defective AKT-Miro1 interaction observed in insulin-resistant states may contribute to mitochondrial dysfunction by preventing appropriate mitochondrial adaptation to insulin signaling, thereby exacerbating insulin resistance.

Our findings demonstrate that exercise training alleviates skeletal muscle mitochondrial hyper-fragmentation and Miro1 in patients with obesity and T2D, which was associated with improved mitochondrial function and insulin sensitivity. These results align with previous studies showing that exercise reduces excessive mitochondrial fission through the AMPK/Irisin pathway, improving glucose metabolism and inflammation.^37^ The observed reductions in DRP1 phosphorylation, Mid49, and Mid51 expression suggest that exercise promotes a shift toward mitochondrial elongation, consistent with reports that endurance training enhances mitochondrial connectivity and efficiency.^26,38–42^ Notably, despite significant changes in mitochondrial morphology, we found no differences in PGC1α, MFN1, MFN2, OPA1, PINK1, or Parkin expression, suggesting that exercise-induced improvements in mitochondrial function may be mediated more by network remodeling rather than biogenesis. This highlights the importance of mitochondrial dynamics and quality control in metabolic adaptation to exercise. The increase in OXPHOS capacity and respiratory conductance further supports the role of mitochondrial remodeling in enhancing energy efficiency, as previously reported in both human and animal models.^43–45^ Additionally, the reduction in LC3 expression suggests a decrease in mitophagy signaling, potentially indicating enhanced mitochondrial quality that reduces the need for turnover. Given that mitochondrial dysfunction and impaired trafficking contribute to insulin resistance, the exercise-induced reduction in Miro1 accumulation suggests a previously underappreciated role for Miro1 in skeletal muscle insulin signaling. These findings reinforce the therapeutic potential of exercise in restoring mitochondrial function and metabolic health in T2D, providing further evidence that targeting mitochondrial dynamics may be an effective strategy for improving insulin resistance. Some limitations are noted. The number and prescription of glucose lowering medications was broad. Thus, the variability in response to exercise training may be explained, in part, by medication use. Furthermore, few patients were using incretin analogues, which may limit generalizability within the landscape of current diabetes care. Thus, further research is required to determine whether modulating Miro1 directly alters muscle contractility or responsiveness to exercise training.

Our study demonstrates that skeletal muscle-specific Miro1 deletion enhances insulin sensitivity and mitochondrial oxidative phosphorylation, supporting a direct, non-canonical role for Miro1 in regulating glucose metabolism. These findings align with prior work linking mitochondrial dynamics and insulin action, where excessive mitochondrial fragmentation has been implicated in insulin resistance.^14^ The observed increase in AKT and AS160 phosphorylation, along with enhanced GLUT4 translocation in Miro1^SkM-/-^ mice, suggests that Miro1 deletion potentiates insulin signaling, leading to improved glucose uptake. This contrasts with previous reports where defects in mitochondrial trafficking and turnover were associated with metabolic dysfunction,^46^ indicating that the impact of Miro1 on metabolism may be tissue- and context-dependent. Furthermore, the increase in OXPHOS capacity and electron transport chain (ETC) efficiency in Miro1- deficient muscle suggests that Miro1 constrains mitochondrial respiratory capacity. Previous studies have shown that mitochondrial quality control mechanisms, including mitophagy, are essential for maintaining oxidative capacity and insulin sensitivity.^47^ Our findings extend this notion by suggesting that excessive Miro1 accumulation may disrupt these processes, leading to metabolic impairments. The congruence between our in vivo and in vitro results further supports a muscle-autonomous role for Miro1 in glucoregulatory function, independent of systemic adaptations.

Taken together, our findings establish skeletal muscle Miro1 accumulation as a hallmark of insulin resistance in the context of T2D. We demonstrate that impaired mitochondrial AKT translocation in response to insulin underlies Miro1 accumulation, contributing to disrupted glucoregulatory function in skeletal muscle. Notably, exercise training reverses these deficits by promoting mitochondrial network remodeling, improving oxidative phosphorylation, and restoring insulin sensitivity. Furthermore, skeletal muscle-specific deletion of Miro1 enhances glucose uptake and bioenergetic function, confirming its role as a negative regulator of insulin action. Collectively, these data position Miro1 as a novel link between mitochondrial quality control and insulin resistance, highlighting its potential as a therapeutic target for T2D.

## METHODS

### EXPERIMENTAL MODELS AND STUDY PARTICIPANT DETAILS

#### Human Study Participants

##### Human Study 1: Cross-sectional assessment of healthy, overweight/obesity, or type 2 diabetes

In a cross-sectional design, adults between the age of 18-60 were prospectively assigned to one of three groups: healthy weight (HW; BMI 18-24.9 kg/m^2^ without T2D), obesity (Ob; BMI: 30-50 kg/m^2^ without T2D), or obesity with type 2 diabetes (T2D; BMI 30-50 kg/m^2^ with T2D) (**Methods S1**). Participants assigned to T2D had a BMI ≥30 kg/m2 and either had a prior diagnosis of T2D or met American Diabetes Association criteria for diagnosis upon screening by a physician.^49^ Following screening, participants were admitted for a three-day inpatient period during which they were provided standardized meals (55% carbohydrate, 35% fat, and 10% protein) based on individual energy needs. On the morning of day 2, participants were assessed for body composition, followed by an incremental aerobic capacity test. Participants were discharged following completion of testing and readmitted at 1700h for an additional overnight stay and dietary control period. At 2000h, patients were placed on NPO status with exception to water for the remainder of the study. On the morning of day 3, patients were awoken at 0530h for catheterization, followed by initiation of a glucose tracer at 0600h, a muscle biopsy at 0700h, and initiation of single-stage hyperinsulinemic-euglycemic clamp study at 0800h. The study was approved by the Institutional Review Boards of the Cleveland Clinic and Pennington Biomedical Research Center, and all participants provided informed consent. Data were collected between April 2016 and July 2023. A sub-group of patients from this prospective observational study concomitantly participated in a trial registered on clinicaltrials.gov (NCT02697201 and NCT02977442, respectively).

##### Human Study #2: A Randomized Controlled Trial of Exercise Training in Patients with Obesity and Type 2 Diabetes

The Dynamics of Muscle Mitochondria in Type 2 Diabetes (DYNAMMO-T2D) study was a parallel group randomized controlled trial to assess the effects of exercise training on mitochondrial dynamics in adults with obesity and type 2 diabetes (**Methods S2**). Sedentary adults ages 18-60 yrs with obesity (BMI ≥ 30 kg/m^2^) and T2D (HbA1c ≥ 6.5%) were randomized in blocks of 4 (1:1) by a blinded biostatistician to 12 weeks of American Diabetes Association (ADA) Standards of Care in Diabetes^50^ (Ctrl) or aerobic exercise training (Ex). Patients randomized to Ctrl continued with physician-guided medical therapy for 12 weeks in conjunction to receiving American College of Sports Medicine guidelines for exercise and nutritional therapy for T2D.^51^ Patients randomized to Ex underwent 12 weeks of supervised aerobic exercise training for 5 days per week, 1 hour per day, at ∼85% of HRMAX (∼70%of VO2MAX).^52^ Immediately prior to and upon completion of the 12-week study, participants were admitted for a 3-day inpatient stay consisting of body composition, aerobic capacity, and insulin sensitivity testing, as well as undergoing muscle biopsy procedures. During the three-day inpatient period during which they were provided standardized meals (55% carbohydrate, 35% fat, and 10% protein) based on individual energy needs. On the morning of day 2, participants were assessed for body composition, followed by an incremental aerobic capacity test. Participants were discharged following completion of testing and readmitted at 1700h for an additional overnight stay and dietary control period. At 2000h, patients were placed on NPO status with exception to water for the remainder of study. On the morning of day 3, patients were awoken at 0530h for catheterization, followed by initiation of a glucose tracer at 0600h, a muscle biopsy at 0700h, and initiation of the two-stage hyperinsulinemic-euglycemic clamp study at 0800h. The study was approved by the Institutional Review Boards at the Cleveland Clinic and Pennington Biomedical Research Center, and patients provided written informed consent before participation. Data were collected between April 2016 and November 2023. The *Dynamics of Muscle Mitochondria in Type 2 Diabetes Exercise* (DYNAMMO-T2DEX) trial was registered on clinicaltrials.gov (NCT02977442) on 11/28/2016, prior to the enrollment of study participants. Inclusionary criteria were expanded in 2020 allowing for increased BMI (from ≤40 to ≤50 kg/m^2^) and use of hypoglycemic medications to improve recruitment and comply with changes to standards of care for type 2 diabetes. The a priori power analysis indicated a target sample size of N=60 to detect a 28.4% difference in DRP1 phosphorylation with 80% power. In 2021, interim analysis was conducted, and sample size estimates were reduced to N=24 due to increased power (critical t = 2.02 at actual power of 0.85). Neither the patients nor the public were involved in the design, conduct, and reporting of the trial. Enrolled patients did not have access to the random allocation sequence but were open to the study allocation.

#### Animal Models

##### Mouse Study #1: Diet-Induced Obesity and Hyperglycemia (FFClow vs FFChigh)

18-week-old male C57BL/6J mice were randomized to either a low fat (10%), fructose (0%), and cholesterol (0%) (FFClow) or a high fat (40%), high fructose (20%), high cholesterol (2%) diet (FFChigh) for 28 weeks. Body weight and food intake were measured weekly, and body composition was measured every four weeks by NMR (Bruker LF110). After 28 weeks of treatment, mice were euthanized by inhalation of isoflurane and cervical dislocation, and blood and tissues were collected.

##### Mouse Study #2: Genetically Induced Hyperglycemia (Wild-type vs A^y^; Lepr^db/+^ vs _Lepr_db/db_)_

Male wild-type C57BL/6 mice (WT) and mice with the *Lethal yellow* mutation (A^y^) (n=8/genotype) were housed under standard housing conditions (21-23°C) and fed a chow diet (Purina 5015). Body weight and food intake were measured weekly, and intraperitoneal glucose tolerance was assessed at 14 weeks of age. At 16 weeks of age, mice were euthanized by carbon dioxide inhalation and cervical dislocation, and blood and tissues were collected.

Additionally, quadricep muscles from 10-week-old male Lepr^db^ heterozygous (db/+) and knockout (db/db) mice (n=8/genotype) were purchased from the Jackson Laboratory.

##### Mouse Study #3: Ex-vivo Insulin Signaling in HFD

Male C57BL/6 mice were bred in-house and assigned to a high fat (60%) diet (HFD) at 14 weeks of age. At 30 weeks of age, mice were euthanized by inhalation of isoflurane and cervical dislocation, and blood and tissues were collected. Gastrocnemius muscles were exposed to insulin ex-vivo as described in detail below.

*Mouse Study #4: Miro1^fl/fl^ vs Miro1^SkM-/-^*

14-week-old male mice with a skeletal muscle-specific deletion of Miro1 (Miro1^SkM-/-^) mice and littermate controls (Miro1^fl/fl^) were assigned to HFD for 16 weeks. Body weight and food intake were measured weekly, and body composition was assessed monthly. Intraperitoneal insulin tolerance tests (ITT) were performed at 28 weeks of age (14 weeks of treatment). At 30 weeks of age (16 weeks of treatment), mice were euthanized by inhalation of isoflurane and cervical dislocation, and blood and tissues were collected.

#### Cell Culture and Lines

C2C12 cells (passage 2) were purchased from American Type Culture Collection and expanded in Dulbecco’s modified Eagle medium (DMEM) supplemented with 4.5 g/l D-glucose (25mM), 50U/ml penicillin/streptomycin, and 10% fetal bovine serum. Cells were incubated at 37°C, 5% CO2 in a humidified incubator. After reaching 100% confluence, cells were induced to differentiate by replacing the growth medium with high-glucose DMEM supplemented with 2% horse serum and allowing cells to differentiate for 48 h unless otherwise indicated. In both growth and differentiation stages, the medium was replenished daily.

### METHOD DETAILS

#### Reagents and Resources

All antibodies, chemicals, reagents, animal diets, and software are listed in detail in Supplementary Table 1.

#### Human Studies

##### 3-day Inpatient Stay

Participants arrived in the evening on day 1 of the inpatient stay and were provided a standardized meal consisting of 55% carbohydrate, 35% fat, and 10% protein with caloric composition based on individual needs. Body composition and incremental aerobic capacity tests were performed on the morning of day 2. After completion of testing, participants were discharged and returned at 1700 for another overnight stay and dietary control period identical to day 1. Patients were placed on NPO except for water at 2000 for the remainder of the inpatient stay. Patients were awoken at 0500 on the morning of day 3 for catheterization. Glucose tracer was initiated at 0630, and a muscle biopsy was collected at 0700. The hyperinsulinemic-euglycemic clamp was initiated at 0800. A second muscle biopsy was collected at 1100 under insulin-stimulated conditions. Baseline and insulin-stimulated whole body respiratory exchange ratios (RER) and substrate metabolism were measured via indirect calorimetry.

##### Body Composition

Body composition was measured as previously described.^53^ Briefly, height and weight were measured in a pre-weighed hospital gown. Dual-energy X-ray absorptiometry (Lunar iDXA, Madison, WI, USA) was used to determine whole body fat and lean mass. iDXA software was used to estimate fat and lean mass according to manufacturer instructions.

##### OGTT

Glucose tolerance and insulin secretion were determined by an oral glucose tolerance test (OGTT) as described previously.^54^ After an overnight fast, patients were given a 75g glucose challenge. Blood samples were obtained from an indwelling intravenous line at baseline (0), 30, 60, 90, 120, and 180 minutes. Samples were immediately assayed for glucose (YSI 2300; STAT Plus, Yellow Springs, OH, USA), and plasma was isolated by centrifugation at 1000 rpm for 10 min at 4°C and stored at -80°C until time to assay for insulin and C-peptide. Calculations for total area of the curve of glucose (tAUC glucose), total area of the curve of insulin (tAUC insulin), total area of the curve of c-peptide (tAUC c-peptide), homeostatic model of insulin resistance (HOMA-IR), disposition index, and Matsuda index were made from equations described previously.^55^

##### Blood Chemistry

Metabolic profiles were analyzed using an automated platform as described.^52^ Cell counts and lipid concentrations were included in the automated blood chemistry platform. Insulin was determined by a commercially available ELISA.

##### Aerobic Capacity

Maximal oxygen consumption (VO2max) was determined using an incremental, graded treadmill test as described previously.^42^ The following criteria were used to determine a maximal test: 1) oxygen consumption plateau (<150 ml/min), 2) heart rate within 15 beats of age-predicted maximum, 3) RER > 1.15, and/or 4) volitional fatigue. Three of the four criteria were required to be met for the test to be considered maximal.

##### Insulin Sensitivity

Insulin sensitivity was determined using a two-stage hyperinsulinemic-euglycemic clamp (90 mg/dl, 40 mU·m^−2^·min^−1^). Medications for T2D were withheld for at least 48 hours prior to the hyperinsulinemic-euglycemic clamp. A constant infusion of insulin and variable infusion of 20% dextrose began at 0 min. Venous blood was sampled every 5 min from an arterialized heated hand to measure glucose (YSI 2900 Biochemistry Analyzer, YSI, Inc., Yellow Springs, OH, USA). The glucose infusion rate (GIR) was adjusted accordingly to maintain plasma glucose at 90 mg/dL. Insulin sensitivity was calculated as insulin-stimulated glucose metabolism (M; mg·kg^−1^·min^−1^) divided by plasma insulin (I; μU/ml) over a 30-minute steady-state period. Plasma for assessing glucose kinetics was deproteinized, extracted, and derivatized before analysis by gas chromatography-mass spectrometry. Isotopic enrichment was determined by fitting the fractional abundances (M+2; m/z 330)/(M0; m/z 328) against a calibration curve. The rate of glucose appearance was then derived using the Steele equation.^56^ Whole body RERs and substrate metabolism were determined under basal and insulin-stimulated conditions via indirect calorimetry (Vmax Encore, Viasys, Yorba Linda, CA, USA).

##### Exercise Training RCT: Standard Care Group

Patients randomized to Ctrl received standard medical therapy throughout the duration of the study. Patients continued all prescribed medications and were given lifestyle and diet recommendations based on ADA guidelines as prescribed by the patient’s healthcare provider.

##### Exercise Training RCT: Exercise Training Group

Exercise consisted of walking and running on a treadmill or stationary cycling on a cycle ergometer as previously described.^52^ Participants completed exercise sessions 5 days/week for 50-60 min/session for 12 weeks. Initial exercise prescription was 55-60% of heart rate max (HRmax) and gradually increased so that after 2-3 weeks, participants were exercising at 80-85% HRmax (∼70% VO2max). All exercise sessions were supervised by an Exercise Physiologist and were conducted in either the Walker Fitness Center at the Cleveland Clinic or at the Fitness Center at Pennington Biomedical. Heart rate monitors (Polar Electro Inc., Woodbury, NY, USA) were worn during exercise to provide visual feedback of individual target heart rates. Compliance with training was documented through regular attendance at the training sessions, heart rate recording, and the use of training logs. Dietary intake was monitored by having each subject complete 3-day food records at baseline and at 4-week intervals during the intervention. Calorie and macronutrient intake data were used as covariates.

##### Skeletal Muscle Biopsy

Skeletal muscle specimens were collected from the *vastus lateralis* using a modified Bergström biopsy technique following an overnight fast as described previously^57^ prior to the start of the hyperinsulinemic-euglycemic clamp. Upon collection, fat and connective tissue were dissected. Samples were immediately placed into preservation media or frozen in liquid nitrogen. Flash-frozen samples were stored at −140°C until the time of analysis.

##### Mitochondrial Isolation

Mitochondria were isolated from fresh skeletal muscle tissue as previously described.^58,59^ Skeletal muscle tissue sectioned at time of biopsy was immediately placed on ice-cold BIOPS preservation buffer (50 mM K+-MES, 20 mM taurine, 0.5 mM dithiothreitol, 6.56 mM MgCl2, 5.77 mM ATP, 15 mM phosphocreatine, 20 mM imidazole, pH 7.1, adjusted with 5M KOH at 0°C, 10 mM Ca–EGTA buffer, 2.77 mM CaK2EGTA + 7.23 mM K2EGTA; 0.1 mM free calcium) until processing. Fresh tissue was transferred to mitochondrial isolation buffer (200 mM sucrose, 10 mM Tris, 0.1 M EGTA/Tris) in the presence of protease and phosphatase inhibitors and minced with scissors on ice. Tissue pieces were transferred to a 7mL glass-on-glass dounce homogenizer with 1.5 mL of mitochondrial isolation buffer and homogenized for 12 strokes. Homogenate was transferred to a 1.7 mL Eppendorf tube and centrifuged at 600 x *g* for 10 minutes at 4°C. The supernatant was collected and centrifuged at 7,000 x *g* for 10 minutes at 4°C. The resulting supernatant (cytosolic fraction) was collected and stored at -80°C until further analysis. The remaining mitochondrial pellet was cleaned with 1.5 mL of mitochondrial isolation buffer and centrifuged again at 7,000 x *g* for 10 minutes at 4°C. The supernatant was removed, and the mitochondrial pellet was incubated on ice until analysis of respiratory function. Following respiratory assays, the remaining mitochondria were re-pelleted and stored at -80°C until further analysis.

##### Skeletal Muscle OXPHOS and ET Capacity

Skeletal muscle OXPHOS and ET capacity were determined *ex vivo* in isolated mitochondria as described previously.^13^ Freshly prepared mitochondrial pellets were resuspended in Buffer D (105 mM KMES, 30 mM KCl, 10 mM KH2PO4, 5 mM MgCl2, 1 mM EGTA, with 2.5 g/L of defatted BSA, pH 7.2)^28^ and injected into an oxygraphy chamber (∼150mg) containing 2 mL of Buffer D. OXPHOS and ET capacity were determined using the following concentrations of substrates, uncouplers, and inhibitors: malate (2 mM), pyruvate (2.5 mM), ADP (2.5 mM), glutamate (10 mM), succinate (10 mM), palmitoylcarnitine (10 μM), duroquinol (0.5 mM), tetramethyl-p-phenylenediamine (TMPD, 0.5 μM), ascorbate (2 mM), carbonylcyanide-p-trifluoromethoxyphenylhydrazone (FCCP, 0.5 μM increment), rotenone (75 nM), antimycin A (125 nM) and sodium azide (200 mM). Cytochrome c was added to confirm membrane integrity. Oxygen flux was normalized to mitochondrial protein content measured by BCA.

##### Skeletal Muscle OXPHOS Conductance

Skeletal muscle OXPHOS conductance was assessed in isolated mitochondria by the creatine kinase clamp technique as previously described with minor modifications.^28^ Briefly, isolated mitochondria were resuspended in Buffer D and injected into an oxygraph chamber with Buffer D, ATP (5 mM), creatine (5 mM), phosphocreatine (1 mM), creatine kinase (200 units/mL). Substrate oxidation was supported by either glutamate and malate to derive NADH-linked conductivity or succinate and rotenone to derive succinate-linked conductivity. The extramitochondrial ATP:ADP ratio was then adjusted by sequential titration of phosphocreatine (3-30mM). OXPHOS capacity was measured in the presence of saturating substrate. To determine conductance, the ionic strength of the Buffer D was calculated and ΔG of ATP hydrolysis at each addition of phosphocreatine. Steady-state mitochondrial respiration at each stage was plotted against ΔG of ATP hydrolysis, and a linear regression was used to determine the slope, which is ∼1/R = conductance. The extramitochondrial ATP:ADP ratio was then adjusted by sequential titration of phosphocreatine (3-30 mM) was then titrated from 1 to 30 mM. OXPHOS capacity was derived as the supported in the presence of saturating substrate. To determine conductance, the ionic strength of the Buffer D was calculated, and ΔG of ATP hydrolysis at each addition of phosphocreatine was determined. Steady-state mitochondrial respiration at each stage was plotted against ΔG of ATP hydrolysis, and a linear regression was used to determine the slope, which is ∼1/R = conductance.

##### Western Blotting

Protein was extracted from skeletal muscle tissue and Western blotting was performed as previously described.^42^ Briefly, frozen skeletal muscle specimens were homogenized in ice-cold cell extraction buffer with added protease inhibitor cocktail, 5 mM phenylmethylsulfonyl fluoride, 1 mM sodium orthovanadate and PhosSTOP using a Polytron. Homogenates were incubated on ice for 30 min and centrifuged for 10 min at 14,000 rpm at 4°C. The supernatant was collected, and protein content was measured using a BCA protein assay kit. 2 µg/µL of lysate were solubilized in Laemmli buffer containing 5% β-mercaptoethanol and boiled for 5 min. Samples were loaded into tris glycine gels and separated via SDS electrophoresis. Gels were transferred to PVDF membranes and blocked with 3% non-fat dry milk in tris-buffered saline with 0.1% Tween- 20 (TBST) for 1 hr. Membranes were incubated overnight with primary antibodies (**Table S1**), washed with TBST, and incubated with species-specific horseradish peroxidase-conjugated secondary antibodies (**Table S1**). Immunoreactive proteins were visualized by enhanced chemiluminescence reagent and quantified by densitometric analysis using ImageJ. Visible bands reactive against an internal control were subject to quantification. Gel-to-gel variation and equal protein loading were controlled using a standardized sample on each gel and values were normalized to loading control (β-actin) and expressed as relative change from baseline, unless otherwise stated.

##### Skeletal Muscle Lipid Droplet and Contractile Structure

Intramuscular lipid and myofibril density were spatially resolved and quantified by Focused ion beam scanning electron microscopy (FIB-SEM) and segmentation analysis as described previously (**Methods S3**).^60^ ∼25 mg of muscle tissue was collected at 4% paraformaldehyde, 2% glutaraldehyde in 0.1 M phosphate buffer, pH 7.2. Samples were cut into 1 mm sections and fixed 2.5% glutaraldehyde, 1% paraformaldehyde and 0.12 M sodium cacodylate solution, pH 7.2 then transferred to a 4% osmium solution. Samples were stained with 2% osmium and incubated overnight in a 1% uranyl acetate solution at 4°C. Samples were washed with water and dehydrated with increasing concentrations of ethanol, then incubated in a 50% Epon: 50% ethanol solution for 4 hours in a vacuum sealed container followed by 75% Epon, 25% ethanol solution overnight. The last day of sample preparation consisted of three 100% Epon resin incubations. Samples were place onto aluminum SEM mounts (Carl Zeiss Microscopy GmbH, Jena, Germany) after removing excess resin and placed in a 60°C oven for 48 hours to polymerize. Sample stubs were then mounted in a UCT Ultramicrotome and faced with a Trimtool 45 diamond knife at a thickness feed of 100 nm and a rate of 80 mm/s. Images were acquired with a Crossbeam 540 using Atlas 5 software. Lipid droplets were segmented and quantified using the deep learning model called MitoNet.^48^ Myofibrils were segmented and quantified as described previously.^61^

##### Mitochondrial Ultrastructure and Content

Two dimensional mitochondrial structures were resolved using transmission electron microscopy as previously described.^62^ Briefly, 15–20 mgs of muscle tissue were fixed by immersion in a triple aldehyde-DMSO mixture.^63^ Tissue blocks were post-fixed in ferrocyanide-reduced osmium tetroxide, soaked in acidified uranyl acetate, dehydrated in ascending ethanol concentrations, passed through propylene oxide, and embedded in Poly/Bed resin. Thin sections were stained with acidified uranyl acetate^64^ followed by modified Sato’s triple lead stain.^65^ Mitochondrial elongation was determined deriving the form factor from manually traced, clearly discernible outlines of mitochondria using the mitochondrial shape descriptors plugin in ImageJ 4 as described previously.^66^

#### Animal Studies

##### Animal Acquisition, Housing, and Husbandry

###### Mouse Study 1: Diet-Induced Hyperglycemia (FFClow vs FFChigh)

Male C57BL/6 mice were bred in-house and weaned at 3 weeks of age. Mice were multi-housed until the start of experimental diet, at which time they were separated into single-housed cages. Mice were housed at murine thermoneutral (28-30°C) from weaning and for the entire duration of the study. At 18 weeks of age, mice were randomized to a FFClow or a FFChigh diet. Throughout the duration of the diet study, body weight and food intake were measured weekly, and body composition was measured every four weeks by NMR. Mice were euthanized after 28 weeks of experimental diet.

###### Mouse Study 2: Genetically Induced Hyperglycemia (Wild-type vs A^y^; Lepr^db/+^ vs Lepr^db/db^)

Male C57BL/6 and *A^y^* mice were purchased from Jackson Laboratory. Upon arrival, mice were single-housed under standard housing conditions (21-23°C). Mice were fed a standard chow diet. Throughout the duration of the study, body weight was assessed weekly. Mice were euthanized at 16 weeks of age.

Quadricep muscles from 10-week-old db/+ and db/db mice were purchased from Jackson Laboratory. Male heterozygous and *Lepr^db^* mice were bred and maintained by Jackson Laboratory under standard housing conditions and diet. Mice were euthanized by carbon dioxide inhalation at 10 weeks of age, and quadricep tissue was collected and snap-frozen in liquid nitrogen. Tissues were shipped on dry ice and immediately stored at - 80°C upon reception until time of analysis.

###### Mouse Study 3: Ex-vivo insulin signaling in HFD

Male C57BL/6 mice were bred in-house and maintained at thermoneutrality (28-30°C) on a standard chow diet. At 12 weeks of age, mice were placed on a HFD for 16 weeks. After 16 weeks of diet, mice were euthanized. Gastrocnemius muscle was collected into ice-cold BIOPS at the time of necropsy and held on ice until the time of assay (4-6 hrs). Individual gastrocnemius muscles were cleaned of fat and connective tissue and were split into two pieces at the Achille’s tendon. Muscle samples were incubated in DMEM high-glucose media containing 3 µM insulin (Novolin) or DMEM vehicle control for 15 minutes. After incubation, the media was removed, and samples were washed with ice-cold PBS containing 0.1M Na3VO4. Samples in ice-cold PBS were transferred to Eppendorf tubes and centrifuged at 2,000 x *g* for 1 min at 4°C. Following centrifugation, PBS was removed, and samples were immediately snap frozen and stored at -80°C until further analysis.

*Mouse Study 4: Miro1^fl/fl^ vs Miro1^SkM-/-^*

Mice with skeletal muscle-specific deletion of Miro1 were generated and bred in-house. Male Miro1^SkM-/-^ mice and floxed littermate controls were weaned at 3 weeks of age. Mice were multi-housed until the start of experimental diet, at which time they were separated into single-housed cages. Mice were housed at murine thermoneutral (28-30°C) from weaning and for the entire duration of the study. At 14 weeks of age, mice were placed on HFD. Throughout the duration of the diet study, body weight and food intake were measured weekly, and body composition was measured every four weeks by NMR. Mice were euthanized after 16 weeks of experimental diet.

##### Generation and Validation of Mice with Skeletal Muscle-Specific Deletion of Miro1

Mice with flanked *loxP* sites for Miro1 were purchased from Jackson Laboratory. Additionally, transgenic mice with *Cre* recombinase driven by the human Acta1 gene promoter were purchased from Jackson Laboratory (strain #006149) and bred with Miro1- floxed mice to generate a skeletal muscle specific deletion of Miro1. Tail snips were collected from pups at 2 weeks of age for DNA extraction, PCR, and gel electrophoresis to determine genotype. Skeletal muscle specific deletion was confirmed by qPCR and Western blot analysis.

##### DNA Extraction, PCR, and Southern Blotting

Tail snips collected from 2-week-old pups were immediately digested in 500 µL lysis buffer (1M Tris, 5M NaCl, 0.5M EDTA, 10% SDS) in the presence of proteinase K at 55°C overnight. Once digested, 500 µL of phenol-chloroform was added and samples were vortexed and centrifuged for 10 min at 14,000 rpm. The resulting supernatant was centrifuged for 10 min at 14,000 rpm to pellet DNA. Pellets were cleaned with 1 mL of ice-cold 100% ethanol and centrifuged for 10 min at 14,000 rpm. Supernatant was discarded and pellets were dried at room temperature. Pellets were resuspended in TE buffer (1M Tris, 0.5M EDTA) and stored at 4°C until further analysis. PCR was performed using GoTaq® Flexi DNA Polymerase kit using 50-100 ng of DNA in the presence of primers for either the Acta1 *cre* or the Miro1 *loxP* sites based on sequences provided by Jackson Laboratory (**Table S2**). Amplification of *cre* and *loxP* DNA was visualized using 1.5-2.5% agarose gels.

##### Intraperitoneal Insulin Tolerance Test

Insulin tolerance tests (ITT) were performed as described previously with minor modifications.^67^ Briefly, mice were fasted for 3 hrs prior to the start of the ITT and body weights were obtained. The tail was cleaned with a 70% isopropanol wipe before creating a tail nick with a lancet. Baseline blood glucose was measured beginning at 30 minutes prior to the insulin injection using a glucometer. A 1 IU/kg dose of insulin in saline was injected into the peritoneum at timepoint 0. Blood glucose was measured by reopening the scab from the lancet nick at 10, 20, 30, 45, 60, 75, 90, and 120 min after insulin injection, and area under the curve was calculated for each mouse.

##### Skeletal Muscle ex vivo Insulin Signaling

Gastrocnemius muscle was collected into ice-cold BIOPS at time of necropsy and incubated in DMEM high-glucose media containing 3 µM insulin or DMEM vehicle control for 15 minutes as described above.

##### Cellular Fractionation

Mitochondria and cytosol were fractioned from frozen gastrocnemius tissue as described above. The cellular membrane was fractioned from the resulting pellet containing the nuclear and membrane fractions from the first centrifugation of the isolation using a commercially available cell membrane extraction kit. Nuclear pellets were resuspended in 1 mL cytosolic buffer (add company/recipe) in the presence of protease and phosphatase inhibitors, then briefly vortexed and sonicated. Samples were centrifuged for 5 min at 500 x *g* at 4°C. Resulting supernatant was discarded and membrane buffer plus protease and phosphatase inhibitors were added to pellets. Samples were briefly vortexed and sonicated again and were incubated on a rotary tube revolver at 4°C for 30 min. Samples were centrifuged for at 3,000 x *g* for 10 min at 4°C. Resulting supernatant containing the membrane fraction was collected. Protein content of each subcellular fraction was measured using BCA.

##### Western Blotting

1-3 µg/µL of mitochondrial, cytosolic, and nuclear lysate were used for Western blotting as described above. Membranes were incubated overnight with primary antibodies (**Table S1**) and visualized using chemiluminescence as described above.

##### Co-Immunoprecipitation

AKT antibody was precipitated by incubating 10 µg of mitochondrial protein with protein A/G agarose beads at 1:10 dilution overnight at 4°C as previously described.^53^ Protein was pulled down by centrifuging at 1000 rpm for 1 min at 4°C. Pellets were washed and then denatured by boiling for 5 min in the presence of 5x SDS sample buffer. Samples were mixed and centrifuged at 10,000 x *g* for 3 min at 4°C. Resulting supernatant was loaded into tris glycine gels for electrophoresis and Western blotting as described against Miro1. Horseradish peroxidase–tagged secondary antibody was used. Input protein, negative beads-only control, and IgG controls were also blotted.

##### Skeletal Muscle OXPHOS and ET Capacity

Skeletal muscle OXPHOS and ET capacity were measured as described above using isolated mitochondria from quadricep muscle. Mitochondria were isolated from fresh quadricep muscle collected into ice-cold BIOPS at time of necropsy as described above. Mitochondrial protein concentrations were determined by BCA. Mitochondrial pellets were resuspended in ice-cold Buffer D and 20 µg of mitochondrial protein were loaded into 2 mL oxygraphy chambers containing Buffer D.

##### Skeletal Muscle Mitochondrial Conductance

Mitochondrial conductance was determined by creatine kinase clamp using isolated mitochondria from quadricep muscle as described above. Mitochondrial pellets were resuspended in ice-cold Buffer D and 20 µg of mitochondrial protein were loaded into 2 mL oxygraphy chambers containing Buffer D supplemented with ATP, creatine, phosphocreatine, and creatine kinase.

#### Cell Culture and Lines

##### Miro1 Deficient C2C12 Cells

Transgenic C2C12 cells were generated by expressing a lentiviral construct targeting pLKO.1 or shMiro1 at a multiplicity of infection of 50. Lentiviral particles were added to cell culture media enriched with 0.5 μg/mL polybrene, added to cells dropwise, and incubated for 24 hours. This process was repeated for 2 days, followed by puromycin selection (2 μg/mL) for 48 hours. Transfection efficiency was confirmed by qPCR and Western blot. Cell experimentation was conducted on passages 3-7.

##### AKT Inhibition

C2C12 cells at passage 4 were grown to 90% confluency in high glucose DMEM supplemented with 10% FBS and 1% Penicillin Streptomycin. At 90% confluency, cells were washed with warm PBS and incubated with 5µM MK2206 dihydrochloride in high glucose DMEM for 2 hours. 15 minutes before cells were collected, 1mM of insulin was added and cells were incubated at 37°C for 15 minutes. Following incubation, cells were washed with ice-cold PBS and collected for pAKT, total AKT, and Miro1 protein expression by Western Blot.

##### Immunofluorescence

Passage 4 C2C12 were grown on 35 mm glass bottom dishes until 50-60% confluency. Cells were washed 1x with warm PBS and incubated in high glucose DMEM with 1mM insulin for 15 minutes at 37°C. Cells were fixed with 4% paraformaldehyde for 30 minutes at room temperature followed by a 20-minute permeabilization with 0.1% Triton X-100 in PBS containing 100 nM glycine. Cells were blocked for 1 hour in 5% goat serum in PBS, followed by overnight primary antibody 1:200 incubation at 4°C. Cells were washed 3x with PBS, then incubated with secondary antibody 1:500 for 2 hours at 4°C. Cells were washed 3x with PBS, and 2mL of PBS was added to the dish with 2 drops of NucBlue Fixed Cell ReadyProbes Reagent (DAPI). Cells were imaged using the Leica DMI6000 B microscope using 40x dry objective lens.

##### Glucose Uptake

Intracellular glucose uptake was determined by radiolabeled glucose uptake assay as described previously with minor modifications.^68^ Briefly, media was aspirated, and cells were gently washed twice with warm PBS then incubated in serum-free low-glucose DMEM supplemented with 2% bovine serum albumin for 4 hours prior to insulin stimulation. Media was aspirated and cells were gently washed twice with warm PBS. Cells were stimulated with vehicle media (Krebs-Ringers-Hepes (KRH) buffer (20 mM Hepes, 136 mM NaCl, 4.7 mM KCl, 1.25 mM MgSO4·7H2O, 1.25 mM CaCl2, pH 7.40) with 2% bovine serum albumin, 2mM Sodium Pyruvate, 32 mM D-Mannitol) or 1 μM insulin in vehicle media for 15 minutes at 37°. After 15 minutes, 25 mM glucose was added to all wells and cells were incubated for an additional 45 minutes. The stimulation medium was then aspirated, and the cells were gently washed twice with warm PBS. Cells were incubated with KRH containing 1 μCi/ml [3H]-2-deoxy-d-glucose and 2 mM 2- deoxy-d-glucose for 10 minutes at 37°. After 10 min of radiolabeled glucose uptake, cells were placed on ice and washed with ice-cold PBS three times and then lysed with 0.1% sodium dodecyl sulfate (SDS) in ddH2O for 30 min with gentle shaking. Lysates were then used for protein quantification and scintillation counting in Ultima Gold scintillation fluid.

##### Cellular Fractionation

C2C12 myotubes were collected in ice-cold mitochondrial buffer plus protease and phosphatase inhibitors immediately following insulin stimulation assay. Mitochondrial, cytosolic, and cellular membrane fractions were obtained as described above. Protein concentrations of each fraction were measured using BCA.

##### Western Blotting

1-3 µg/µL of mitochondrial, cytosolic, and nuclear lysate were used for Western blotting as described above. Membranes were incubated overnight with primary antibodies (see STAR methods) and visualized using chemiluminescence as described above.

##### Quantitative RT-PCR

Cells were washed in ice-cold PBS prior to collection in ice-cold Trizol. RNA was isolated using the RNeasy Mini Kit according to manufacturer’s instructions. CDNA was synthesized using the High-Capacity cDNA Reverse Transcription Kit with RNAse Inhibitor according to manufacturer’s instructions. Knockdown efficiency was validated by quantitative PCR using targeted primers for Miro1:

Forward: CACAAGCTGACCTCAAGAGC Reverse: GCAAAGACCGTAGCACCAAA

##### OXPHOS and ET Capacity

OXPHOS and ET capacity were measured in C2C12 myoblasts as described above. Cells were cultured for 3-4 days until 90-100% confluency was reached as previously described. Cells were briefly washed with PBS, detached using 0.05% trypsin solution, and centrifuged at 600 x *g* for 6.5 min. Cells were resuspended in 2.5 mL of pre-warmed mitochondrial respiration buffer MiR05 (110 mM sucrose, 60 mM K +-lactobionate, 0.5 mM EGTA, 3 mM MgCl2, 20 mM taurine, 10 mM KH2PO4, 20 mM HEPES adjusted to pH 7.1 with KOH at 37°C; and 1 g/L de-fatted BSA) at a final concentration of 5 million cells/mL. 2 mL of cell suspension was added to oxygraphy chambers. Cells were permeabilized using 20 µg/mL digitonin, and maximal OXPHOS and ET capacity were determined as described above.

### QUANTIFICATION AND STATISTICAL ANALYSES

Statistical procedures and corresponding contrasts are detailed in each figure legend. Two- or three-way comparisons of continuous variables were conducted via student’s t-test or by analysis of variance, as appropriate. Abnormally distributed data were compared by non-parametric measures including Mann-Whitney, Tukey’s HSD, and Welch’s ANOVA test, as appropriate based on number of groups. Circumstances with more individualized data handling procedures are detailed below. *<u>Human Study #1</u>*: Between-group differences were evaluated by a one-way ANOVA with 3 group fixed effects. Pairwise comparisons of significant main effects were assessed using Tukey’s HSD test. Distributional assumptions were evaluated using the Brown-Forsythe test. In the case of abnormally distributed data, Welch’s ANOVA test was employed with Dunnett’s T3 test for multiple comparisons. Data were analyzed using Graphpad Prism 10. *<u>Human Study #2</u>*: Differences in baseline characteristics were compared by linear mixed modeling. The influence of covariates on clinical outcomes of interest were evaluated by a type III test of fixed effects. Between-group differences in outcome changes over time following intervention were evaluated by linear mixed modeling. Linear mixed modeling was chosen to account for the hierarchical structure of the data, with repeated measurements nested within individuals, while accommodating missing data under the assumption of missing at random. The primary model included fixed effects for group (intervention vs. control), time (as a categorical variable representing measurement occasions), and the group × time interaction. The interaction term was the primary parameter of interest, testing whether the intervention led to differential changes in the outcome over time relative to the control group. A random intercept for each participant was included to account for individual baseline differences, and a random slope for time was tested to assess individual variation in longitudinal trajectories. Model assumptions, including normality of residuals and homogeneity of variance, were assessed using graphical diagnostics and sensitivity analyses. Robustness checks included alternative covariance structures, the addition of baseline covariates, and multiple imputation for missing data where appropriate. Statistical analysis was conducted using SAS version

9.4. *<u>Lipid Droplet Architecture and Myofibril Assembly</u>:* Linear regression models were used to predict lipid droplet and myofibril characteristics. Individual differences were accounted a covariate model which accounted for essential baseline characteristics (HbA1c, sex, and age). For individual lipid droplet characteristics, statistical significance was set at p = 0.0001 due to the hundreds of lipid droplets per sample. Statistical analysis for the lipid droplet segmentation was performed in R Studio, version 4.1.1.

### DATA AVAILABILITY

This paper does not report the original code. All data are available from the corresponding authors on request. Further information and requests for resources and reagents should be directed to and will be fulfilled by the lead contact, John Kirwan.

## ACKNOWLEDGMENTS

This work was supported by National Institute of Health grants DK108089 (JPK), GM104940 (The Louisiana Clinical and Translational Science Center; JPK), DK072476 (Pennington/Louisiana Nutrition Obesity Research Center), K99AG083239-01 (WSD), AT004094 (ERMZ), GM135002 (Metabolic Basis of Disease Center), DK139728 (ECH), HL169491 (JTM), HL006221 (BG), and HL094307 (PA). The funding bodies had no role in the design of the trial. We thank the Cleveland Clinic and Pennington Biomedical Research Center clinical research units for providing patient care and clinical research support. We thank the study volunteers for their considerable time and effort in the trials. We thank the Comparative Biology Core at Pennington Biomedical Research center for their support with animal care. We acknowledge Randall Mynatt and thank Jaycob Warfel and Allison Stone for sharing data and samples for the A^y^ mouse model. We sincerely acknowledge both past and present members of the IPMM laboratory for their valuable contributions, insightful discussions, and technical support that have helped shape this work.

## AUTHOR CONTRIBUTIONS

Conceptualization: CLA and JPK. Methodology: ALT, BV, HAP, BG, and CLA. Software: RAB and CLA. Validation: ECH, WSD, ALT, BV, RAB, HAP, BG, and CLA. Formal Analysis: ECH, WSD, ALT, HAP, BG, and CLA. Investigation: ECH, WSD, ERMZ, ALT, BV, ERMZ, KPB, JTM, RAB, HAP, BR, CLA, and JPK. Resources: RAB, BG, CLA, and JPK. Data Curation: ECH and CLA. Writing - Original Draft: ECH and CLA. Writing – Review & Editing: ECH, WSD, ERMZ, ALT, BV, ERMZ, KPB, JTM, RAB, HAP, BR, CLA, and JPK. Visualization: ECH, ALT, KPB, HAP, BG, and CLA. Supervision: BG, CLA, and JPK. Project Administration: CLA and JPK. Funding Acquisition: JPK.

