## Supplementary Materials and Methods for "Miro1 Mediates Skeletal Muscle Insulin Resistance in Type 2 Diabetes"

### APPENDIX

**Supplementary Figure 1.** Related to Figure 1. Miro1 accumulation is associated with insulin resistance in patients and mice with obesity and Type 2 Diabetes.

Miro1 expression in heterozygous ($Lepr^{db/+}$) or homozygous ($Lepr^{db/db}$) leptin deficient mice.

**Supplementary Figure 2.** Related to Figure 3. Exercise training relieves skeletal muscle mitochondrial hyper-fragmentation and Miro1 aggregation in patients with obesity and Type 2 Diabetes.

CONSORT diagram depicting participant flow through the randomized controlled trial.

**Supplementary Figure 3.** Related to Figure 3. Exercise training relieves skeletal muscle mitochondrial hyper-fragmentation and Miro1 aggregation in patients with obesity and Type 2 Diabetes.

Ultrastructure analyses of individual skeletal muscle lipid droplet morphology and quantification of lipid droplet content before and after exercise training.

**Supplementary Figure 4.** Related to Figure 3. Exercise training relieves skeletal muscle mitochondrial hyper-fragmentation and Miro1 aggregation in patients with obesity and Type 2 Diabetes.

Expression of proteins related to skeletal muscle mitochondrial biogenesis and turnover.

**Supplementary Figure 5.** Related to Figure 3. Exercise training relieves skeletal muscle mitochondrial hyper-fragmentation and Miro1 aggregation in patients with obesity and Type 2 Diabetes.

NADH-, succinate- and complex III-linked electron transfer (ET) capacity.

**Supplementary Figure 6.** Related to Figure 5. Miro1 negatively regulates skeletal muscle insulin action and oxidative capacity in mice with obesity and hyperglycemia.

Generation of mice with skeletal muscle specific deletion of Miro1.

**Supplementary Figure 7.** Related to Figure 5. Miro1 negatively regulates skeletal muscle insulin action and oxidative capacity in mice with obesity and hyperglycemia.

Effects of skeletal muscle Miro1 deletion on body weight and food intake.

**Supplementary Figure 8.** Related to Figure 6. Miro1 negatively regulates intracellular glucose uptake and oxidative capacity in skeletal muscle cells.

Validation of Miro1-deficent C2C12 cell model.

**Supplementary Table 1.** Key reagents and resources.

**Supplementary Table 2.** Specific western blot antibody conditions.

**Supplementary Table 3.** Primers for genotyping Miro$^{fl/fl}$ and Miro1$^{SkM-/-}$ mice.

**Supplementary Methods 1.** Human Study #1 details: Cross-sectional assessment of healthy, overweight/obesity, or Type 2 Diabetes.

**Supplementary Methods 2.** Human Study #2 details: A randomized controlled trial of exercise training in patients with obesity and Type 2 Diabetes.

**Supplementary Methods 3.** Supplementary experimental methods.

**Supplementary Figure 1.** Related to Figure 1. Miro1 accumulation is associated with insulin resistance in patients and mice with obesity and Type 2 Diabetes.

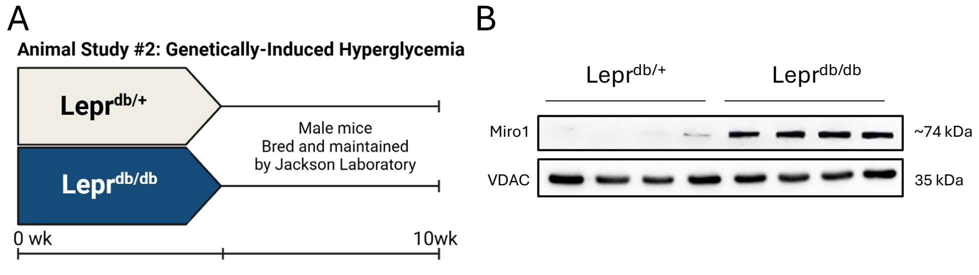


(**A**) Study design of genetically induced hyperglycemia in mice using heterozygous ($Lepr^{db/+}$) or homozygous ($Lepr^{db/db}$) leptin deficient mice (n=4/group). (**B-C**) Western blot and quantification of fold change relative to $Lepr^{db/+}$ for Miro1 expression in skeletal muscle from $Lepr^{db/+}$ and $Lepr^{db/db}$ mice (n=4/group). Data are represented as mean ± SEM. ***p < 0.001 by Student's t test (C).

**Supplementary 2.** Related to Figure 3. Exercise training relieves skeletal muscle mitochondrial hyper-fragmentation and Miro1 aggregation in patients with obesity and Type 2 Diabetes.

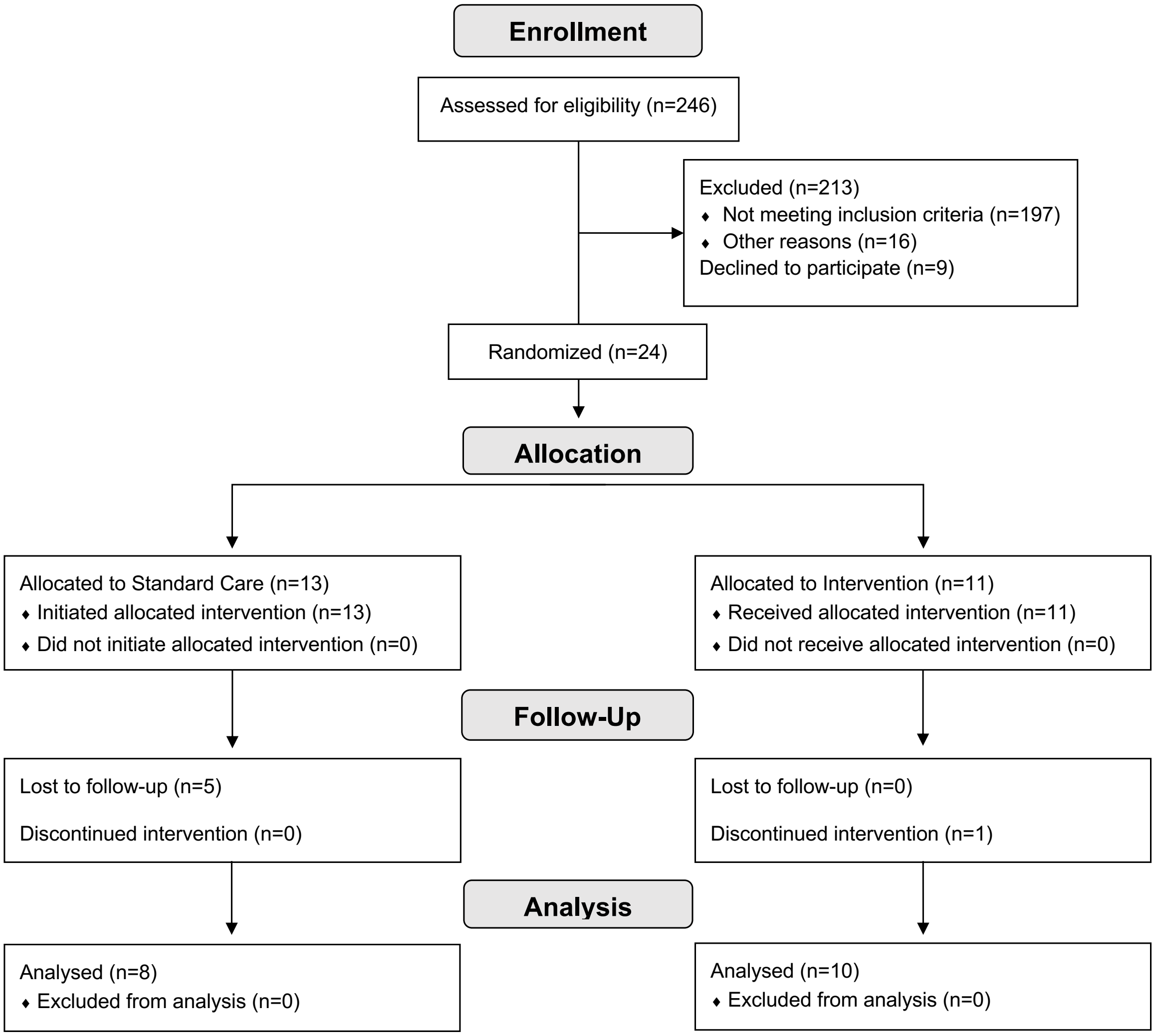


CONSORT diagram illustrating the flow of participants through the DYNAMMO randomized controlled trial (RCT).

**Supplementary Figure 3.** Related to Figure 3. Exercise training relieves skeletal muscle mitochondrial hyper-fragmentation and Miro1 aggregation in patients with obesity and Type 2 Diabetes.

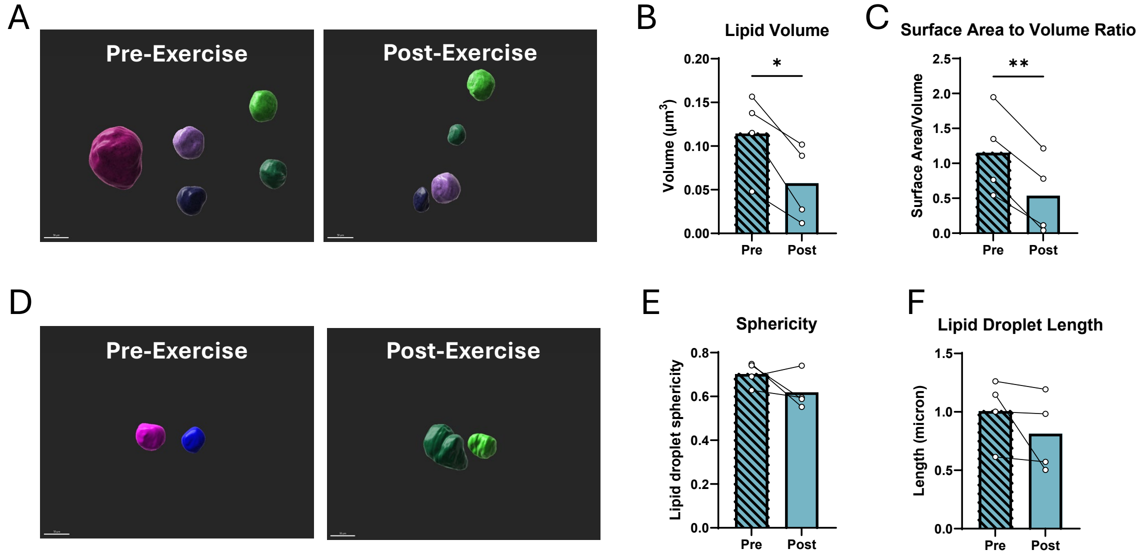


(**A-F**) Ultrastructure analyses of individual skeletal muscle lipid droplet morphology and quantification of lipid droplet content before and after exercise training (n=4). Different colors are representative of individual lipid droplets. Scale bars represent 50μm. Data are represented as mean ± SEM. *p < 0.05, **p < 0.01 by Student's t test (B-C and E-F).

**Supplementary Figure 4.** Related to Figure 3. Exercise training relieves skeletal muscle mitochondrial hyper-fragmentation and Miro1 aggregation in patients with obesity and Type 2 Diabetes.

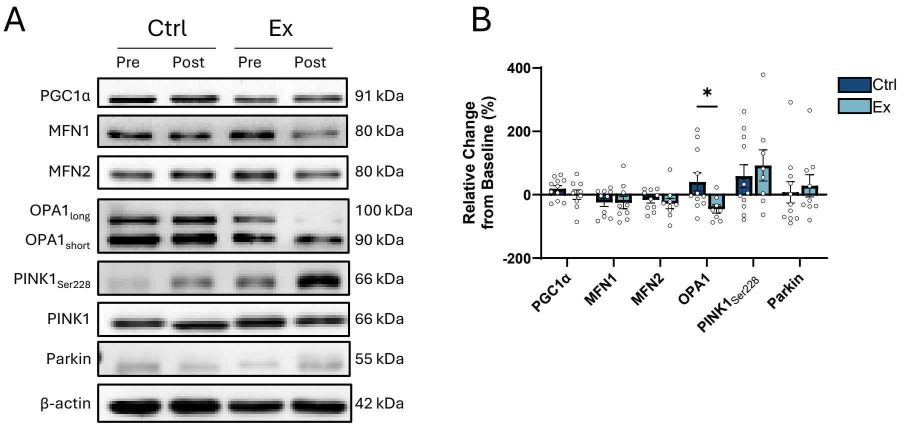


(**A-B**) Western blots and quantification from human skeletal muscle samples in Ctrl and Ex groups before and after 12-week intervention. Relative changes from baseline in PGC1α, MFN1, MFN2, OPA1, PINK1, and Parkin expression before and after 12-week intervention (n=8-12/group). Data are represented as mean ± SEM. *$p < 0.05$ by Student's t test (B).

**Supplementary Figure 5.** Related to Figure 3. Exercise training relieves skeletal muscle mitochondrial hyper-fragmentation and Miro1 aggregation in patients with obesity and Type 2 Diabetes.

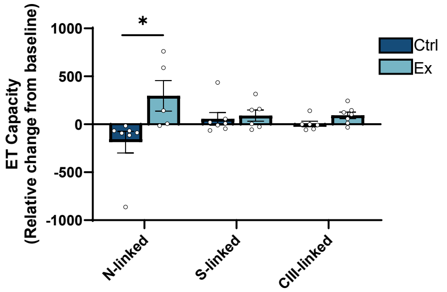


Relative changes from baseline in maximal NADH-linked (N-linked), succinate-linked (S-linked), and complex III-linked (CIII-linked) mitochondrial electron transfer (ET) capacity in Ctrl and Ex patients (n=6-7/group). Data are represented as mean ± SEM and analyzed by Student’s t test.

**Supplementary Figure 6.** Related to Figure 5. Miro1 negatively regulates skeletal muscle insulin action and oxidative capacity in mice with obesity and hyperglycemia.

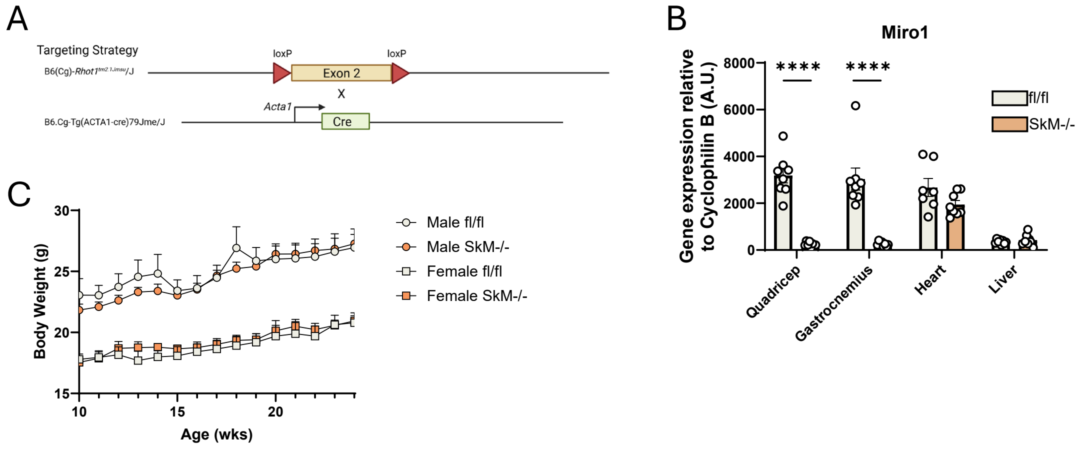


(**A**) Overview of targeting strategy for Miro1 loxP sites and Acta1-Cre promoter. (**B**) Miro1 gene expression across tissues in Miro1 floxed (fl/fl) and mice with skeletal muscle Miro1 deletion (SkM-/-). (**C**) Body weight curves from weekly measurements of Miro1 fl/fl and SkM-/- mice maintained on a low-fat diet. Data are represented as mean ± SEM. ****$p < 0.0001$ by Student's t test (B).

**Supplementary Figure 7.** Related to Figure 5. Miro1 negatively regulates skeletal muscle insulin action and oxidative capacity in mice with obesity and hyperglycemia.

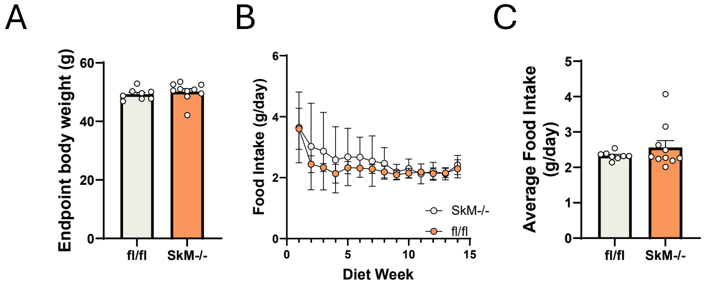


(**A**) Endpoint body weight of Miro1 fl/fl and SkM-/- mice after 16 wks on a high fat diet. (**B**) Weekly food intake curves of fl/fl and SkM-/- mice during experimental diet. (**C**) Calculated average daily food intake of fl/fl and SkM-/- during experimental diet. Data are represented as mean ± SEM and analyzed by Student's t test (A, C)

**Supplementary Figure 8.** Related to Figure 6. Miro1 negatively regulates intracellular glucose uptake and oxidative capacity in skeletal muscle cells.

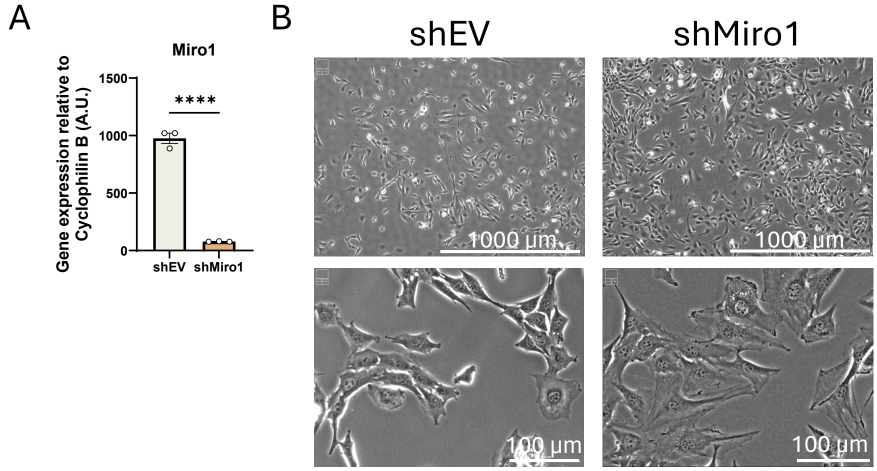


(**A**) Miro1 gene expression in lentiviral empty vector control (shEV) and Miro1 silenced (shMiro1) in C2C12 myoblasts. (**B**) Images of shEV and shMiro1 cells at growth day 4. Data are represented as mean ± SEM. ****p < 0.0001 by Student's t test (A).

| REAGENT or RESOURCE | SOURCE | IDENTIFIER |
|---|---|---|
| **Antibodies** | | |
| RHOT1 (Miro1) | Thermo Fisher | PA5-42646 |
| Phospho-AKT (Thr308) | Cell Signaling | 4056 |
| AKT | Cell Signaling | 9272 |
| GAPDH | Proteintech | 10494-1-AP |
| VDAC | Proteintech | 10866-1-AP |
| Goat anti-rabbit Ig | Millipore | AP132P |
| Veriblot IP detection | Abcam | ab131366 |
| GLUT4 | Abcam | ab33780 |
| Caveolin1 | Cell Signaling | 3267S |
| Phsopho-DRP1 (Ser616) | Cell Signaling | 3455 |
| DRP1 | Cell Signaling | 8570 |
| β-actin | Proteintech | 66009-1-AP |
| Mid49 | Proteintech | 28718-1-AP |
| Mid51 | Proteintech | 20164-1-AP |
| LC3 | Proteintech | 14600-1-AP |
| Phospho-AS160 | Cell Signaling | 2670 |
| TBC1D4 (AS160) | Proteintech | 20024-1-AP |
| PGC1α | Novus | NBP1-04676 |
| MFN1 | Proteintech | 13798-1-AP |
| MFN2 | Cell Signaling | 9482 |
| OPA1 | Proteintech | 27733-1-AP |
| Phospho-PINK1 (Ser228) | Thermo Fisher | PA5-105356 |
| PINK1 | Abcam | ab23707 |
| Parkin | Cell Signaling | 4211 |
| ECL Anti-rabbit IgG | Cytiva | NA9340V |
| ECL Anti-mouse IgG | Cytiva | NA931V |
| TOM20 | Santa Cruz | sc-17764 |
| RHOT1 (Miro1) | Thermo Fisher | PA5-143948 |
| AKT | Proteintech | 60203-2-IG |
| Alexa Fluor 350 | Invitrogen | A11046 |
| Alexa Fluor 488 | Invitrogen | A21131 |
| Alexa Fluor Plus 647 | Invitrogen | A32733 |
| **Bacterial and virus strains** | | |
| pLKO.1-puro Empty Vector Plasmid DNA | Millipore Sigma | SHC001 |
| pLKO.1-puro Rhot1 Bacterial Glycerol Stock | Millipore Sigma | TRCN0000326200 |
| **Biological samples** | | |
| Human vastus lateralis muscle | Self | |
| Human plasma | Self | |
| Human serum | Self | |

| **Chemicals, peptides, and recombinant proteins** | | |
|---|---|---|
| DMEM | Thermo Fisher | 11-885-084 |
| Fetal bovine serum | Cytiva | SH30071.03HI |
| Penicillin-Streptomycin 100x | Sigma Aldrich | P0781-100ML |
| DMEM (high glucose) | Thermo Fisher | 11-995-073 |
| MES hydrate | Sigma Aldrich | M8250-100G |
| Taurine | Sigma Aldrich | T0625-100G |
| Dithiothreitol | Sigma Aldrich | A39255 |
| Magnesium chloride | Millipore Sigma | 7786-30-3 |
| Tris salt of ATP | Millipore Sigma | A9062 |
| Tris salt of phosphocreatine | Millipore Sigma | P1937 |
| Imidazole | Sigma Aldrich | 56750 |
| EGTA | Millipore Sigma | 67-42-5 |
| Calcium chloride | Sigma Aldrich | C5670-100G |
| Sucrose | Sigma Aldrich | S9378-1KG |
| Tris base | Thermo Fisher | BP152-5 |
| Potassium chloride | Sigma Aldrich | P9541-1KG |
| Potassium phosphate monobasic | Sigma Aldrich | 795488-1KG |
| Bovine serum albumin (defatted) | Sigma Aldrich | 03117057001 |
| Sodium pyruvate | Millipore Sigma | 113-24-6 |
| L-Malic acid | Millipore Sigma | 97-67-6 |
| L-Glutamic acid | Millipore Sigma | 142-47-2 |
| Sodium succinate | Millipore Sigma | 6106-21-4 |
| Potassium salt of ADP | Millipore Sigma | 72696-48-1 |
| Creatine monohydrate | Sigma Aldrich | C3630-100G |
| Creatine kinase from rabbit muscle | Millipore Sigma | 10736988001 |
| Cytochrome c from equine heart | Sigma Aldrich | C2506 |
| Duroquinol | TCI | T0822 |
| Carbonyl cyanide 4-(trifluoromethoxy)phenylhydrazone (FCCP) | Millipore Sigma | 370-86-5 |
| Rotenone | Millipore Sigma | 83-79-4 |
| Antimycin A | Millipore Sigma | 1397-94-0 |
| Sodium L-ascorbate | Millipore Sigma | 134-03-2 |
| N,N,N’,N’Tetramethyl-pphenylenediamine dihydrochloride (TMPD) | Thermo Fisher | AAAL00559-09 |
| Sodium Azide | Millipore Sigma | 26628-22-8 |
| Cell extraction buffer | Invitrogen | FNN0011 |
| Protease inhibitor cocktail | Sigma Aldrich | P2714-1BTL |
| PhosSTOP | Roche | 4906837001 |
| Tris glycine gels (4-20%) | Novex | XV04200PK20 |
| Immunoblot PVDF | Bio-Rad | 1620177 |
| Non-fat dry milk | Bio-Rad | 170-6404 |

| | | |
|---|---|---|
| SuperSignal West Pico PLUS chemiluminescent substrate | Thermo Fisher | 34578 |
| Novolin R 100 short-acting insulin | Novo Nordisk | 0169-1833-11 |
| Sodium orthovanadate | Sigma Aldrich | S6508-10G |
| Sodium chloride | Signma Aldrich | S9888-5KG |
| EDTA | Millipore Sigma | 324503 |
| Proteinase K | Thermo Fisher | E00492 |
| GoTaq Flexi DNA Polymerase Kit | Promega | M8291 |
| Mem-PER Plus Membrane Protein Extraction Kit | Thermo Fisher | 89842 |
| Protein A/G Plus-Agarose | Santa Cruz Biotechnology | sc-2003 |
| (H+L) HRP Conjugated | Cytiva | NA9340 |
| MK 2206 dihydrochloride | BioTechne | 7850/10 |
| Polybrene | Sigma Aldrich | TR-1003-G |
| Goat serum | Thermo Fisher | 50197Z |
| Hepes | Sigma Alrich | H4034 |
| Magnesium sulfate heptahydrate | Acros Organics | 423900 |
| D-Mannitol | Sigma Aldrich | M9647 |
| Deoxy-D-glucose, 2-[1,2-3H (N)]- | PerkinElmer | NET328A250UC |
| Ultima Gold scintillation fluid | PerkinElmer | 6013321 |
| Trizol | Invitrogen | 15596026 |
| RNeasy Mini Kit | Qiagen | 74104 |
| cDNA Reverse Transcription Kit with RNAse Inhibitor | Thermo Fisher | 4374966 |
| **Critical commercial assays** | | |
| Human insulin-specific RIA | Millipore Sigma | HI-14K |
| Pierce™ BCA Protein Assay | Thermo Fisher | 23225 |
| **Experimental models: Cell lines** | | |
| C2C12 | ATCC | CRL-1772 |
| **Experimental models: Organisms/strains** | | |
| C57BL/6J | Jackson Laboratories | RRID:IMSR_JAX:000664 |
| B6.BKS(D)-$^{Leprdb/}$J | Jackson Laboratories | RRID:IMSR_JAX:000697 |
| B6(Cg)-Rhot1$^{tm2.1Jmsu}$/J | Jackson Laboratories | RRID:IMSR_JAX:031126 |
| B6.Cg-A$^{y}$/J | Jackson Laboratories | RRID:IMSR_JAX:000021 |
| B6.Cg-Tg(ACTA1-cre)79Jme/J | Jackson Laboratories | RRID:IMSR_JAX:006149 |
| B6(Cg)-Rhot1$^{tm2.1Jmsu}$/J x B6.Cg-Tg(ACTA1-cre)$^{79Jme}$/J | Self | |
| **Software and algorithms** | | |
| Graphpad Prism 10.4.1 | | RRID:SCR_002798 |
| SAS 9.4 | | RRID:SCR_008567 |
| ImageJ 1.54p | | RRID:SCR_003070 |
| MitoNet | Conrad and Narayan[48] | N/A |

| | | |
|---|---|---|
| R Studio, version 4.1.1 | | RRID:SCR_000432 |
| **Animal Diets** | | |
| $FFC_{low}$ (10% fat) | Research Diets | D09100304 |
| $FFC_{high}$ (40% fat, 20% fructose, 2% cholesterol) | Research Diets | D09100310 |
| Standard Chow | Purina | 5015 |
| HFD (60% fat) | Research Diets | D12492 |

**Supplementary Table 1.** Key reagents and resources.

| Primary ab (dilution) | Secondary ab (dilution) |
|---|---|
| RHOT1 (1:1000) | Rabbit (1:8000) |
| Phospho AKT (Thr 308) (1:1000) | Rabbit (1:8000) |
| AKT (1:1000) | Rabbit (1:8000) |
| GAPDH (1:2000) | Rabbit (1:10,000) |
| VDAC (1:4000) | Rabbit (1:10,000) |
| GLUT4 (1:1000) | Rabbit (1:8000) |
| Caveolin1 (1:2000) | Rabbit (1:10,000) |
| Phsopho-DRP1 (Ser616) (1:1000) | Rabbit (1:8000) |
| DRP1 (1:1000) | Rabbit (1:8000) |
| β-actin (1:2000) | Mouse (1:10,000) |
| Mid49 (1:1000) | Rabbit (1:8000) |
| Mid51 (1:1000) | Rabbit (1:8000) |
| LC3 (1:1000) | Rabbit (1:8000) |
| Phospho-AS160 (1:1000) | Rabbit (1:8000) |
| TBC1D4 (AS160) (1:1000) | Rabbit (1:8000) |
| PGC1α (1:1000) | Rabbit (1:8000) |
| MFN1 (1:2000) | Mouse (1:8000) |
| MFN2 (1:3000) | Rabbit (1:8000) |
| OPA1 (1:2000) | Rabbit (1:8000) |
| Phospho-PINK1 (Ser228) (1:1000) | Rabbit (1:8000) |
| PINK1 (1:2000) | Rabbit (1:8000) |
| Parkin (1:2000) | Rabbit (1:8000) |

**Supplementary Table 2.** Specific western blot antibody conditions.

| Primer | FWD | REV |
|---|---|---|
| RHOT1 LoxP | 37011<br>(AAA TGC CAC CAG AAT CCA G) | 37012<br>(GTA GCC CAG TAT GAT GGC ACA) |
| Acta1-Cre | oIMR1084<br>(GCG GTC TGG CAG TAA AAA CTA TC) | oIMR1085<br>(GTG AAA CAG CAT TGC TGT CAC TT) |
| IPC | oIMR7338<br>(CTA GGC CAC AGA ATT GAA AGA TCT) | oIMR7339<br>(GTA GGT GGA AAT TCT AGC ATC ATC C) |

**Supplementary Table 3.** Primers for genotyping Miro$^{fl/fl}$ and Miro1$^{SkM-/-}$ mice.

**Supplementary Methods 1**

Human Study #1: Cross-sectional assessment of healthy, overweight/obesity, or type 2 diabetes

***Inclusion Criteria:***

Group 1 included people who met one of the ADA criteria (FPG ≥ 126 mg/dL, 2-h PG ≥ 200 mg/dL after an oral glucose tolerance test, or HbA1C ≥ 6.5%) consistent with the diagnosis of diabetes and had three or less anti-diabetes medications in their current pharmacotherapy regimen. They were 18 to 45 years of age, sedentary, had a BMI between 25-40 $kg/m^2$, an HbA1C ≤ 9.0%, and had been weight stable for the previous 6 months. Group 2 consisted of age and gender matched, non-diabetic sedentary overweight or obese (BMI, 25-40 $kg/m^2$) volunteers 18 to 45 years of age. Group 3 consisted of age and gender matched, healthy sedentary controls with normal glucose tolerance, a BMI <25 $kg/m^2$, and 18 to 45 years of age. Participants in all groups agreed to allow the collection and storage of specimens, data and images/scans for future use.

***Exclusion Criteria:***

Group 1 patients taking more than three antidiabetic agents or with an $HbA_{1C}$ > 9.0%. Patients with evidence of type 1 diabetes and diabetics requiring insulin therapy, BMI >40 $kg/m^2$, subjects who had not been weight stable (>5kg weight change) in the past 6 months, or subjects who had been recently active (30 min moderate/high intensity exercise two or more times weekly). Tobacco users, smokers or individuals who quit smoking or using tobacco <5 years ago or hypertriglyceridemic (>400 mg/dl) and/or hypercholesterolemic (>260 mg/dl) subjects were excluded. Patients with a history of significant metabolic, cardiac, cerebrovascular, hematological, pulmonary, gastrointestinal, liver, renal, or endocrine disease or cancer that would affect the outcome measures or subject safety were also excluded. Additional exclusion criteria were pregnant (as evidenced by a positive pregnancy test) or nursing women and females who have had a partial or complete hysterectomy. Patients on prescription medications including hormonal contraceptives other than T2D medications or patients requiring regular use of over-the-counter medications that could not discontinue use for study period as required by investigator were excluded. Finally, patients were excluded based on contraindication to exercise.

**Screening**

Screening Visit 1: As part of the first screening visit, each participant reviewed the Informed Consent with a member of the Outpatient Clinic staff. Once the consent was signed, the following were completed: height, weight and vital signs; circumferences measured (waist, hip and neck); medical history and physical; blood work including lipid panels and liver function tests; women had a urine pregnancy test; resting 12 lead ECG; and questionnaires (Minnesota Leisure Time Activity and 3 Day Food Record Instructions).

Screening Visit 2: On the second screening day, the participant reported to the Inpatient Unit to complete a 75g oral glucose tolerance test (OGTT). Blood was collected at specified time points up to 180 minutes post-glucose load, and urine was collected from the start of the OGTT until the 120-minute post-glucose load time point. Patients taking medications for T2D were asked to withhold for at least 48 hours prior to the OGTT. Glucose was evaluated in the evening prior to and the morning of the OGTT. Testing did not proceed and was rescheduled if concentrations >300 or <50 mg/dL were observed. After completing the OGTT, the participant was instructed to maintain current dietary patterns and not initiate a new exercise program. The participant's body weight was used by the research staff to evaluate the maximum allowable blood collection for the remainder of the study. No more than 5 mL/kg of blood in any one 24-hour period or 7 mL/kg in any 8-week period was collected. If the participant exceeded the collection amount described herein, the patient was scheduled accordingly.

**Diet/Weight Stabilization Period and Program Testing**

Participants were required to be weight and diet stable leading into test periods. They had to be stable for at least one week, but timing was adjusted if needed for stabilization. Once it was determined by study staff that the participant was stable and eligible for admission, the participant was admitted to the inpatient unit for testing. Patients were asked to withhold medications for T2D for at least 48 hours prior to the euglycemic clamp test (i.e., the morning of Visit 1). Glucose was evaluated the evening prior to and the morning of the clamp test. Testing did not proceed and was rescheduled if glucose concentrations >300 or <50 mg/dL were observed.

Note: Females had to have their inpatient testing period completed during the mid-follicular phase of the menstrual cycle. Therefore, the timing of the testing period from screening to the first test period was longer if necessary for scheduling purposes. Stabilization was assured as described above.

**Admit Visits**

Admit Visit 1: The participant arrived at the inpatient unit in the late afternoon for an overnight stay for dietary control. Weights and vital signs were collected, along with a completed 3-day food record. Once completed, the participant was provided dinner prior to an overnight fast and was allowed to sleep.

Admit Visit 2: The participant completed a DXA Scan in the morning. Females had a urine pregnancy test prior to the scan. An Aerobic Capacity Test was performed in the Exercise Testing Lab. The participant was allowed to leave the center when these procedures were completed, following the established weight/activity stabilization program. They returned to the inpatient unit at approximately 5 PM for an overnight stay for dietary control. Dinner was provided prior to an overnight fast.

Admit Visit 3: The participant underwent a Euglycemic Clamp Test. Metabolic rate was measured by RMR procedures four times (upon waking and prior to, during, and at the end of the clamp procedure), and two muscle biopsies were completed in conjunction with the clamp procedures. Upon completion of the clamp procedure, the participant was provided a meal and then was discharged from the inpatient unit.

**Supplementary Methods 2**

Human Study #2: A Randomized Controlled Trial of Exercise Training in Patients with Obesity and Type 2 Diabetes

***Inclusion Criteria:***

Participants met one of the ADA criteria (FPG ≥ 126 mg/dL, 2-h PG ≥ 200 mg/dL after an oral glucose tolerance test, or HbA1C ≥ 6.5%) consistent with the diagnosis of diabetes. They were 18 to 60 years of age, sedentary, and had a BMI between 25-50 kg/$m^2$. Subjects agreed to allow the collection and storage of specimens, data, and images/scans for future use.

***Exclusion Criteria:***

Patients with an HbA1C > 10%, evidence of type 1 diabetes and diabetics requiring insulin therapy, or BMI >50 kg/$m^2$ were excluded. Additional exclusion criteria were tobacco users, smokers, or individuals who quit smoking or using tobacco <5 years ago; patients with a history of significant metabolic, cardiac, cerebrovascular, hematological, pulmonary, gastrointestinal, liver, renal, or endocrine disease, or cancer that would have affected the outcome measures or subject safety as determined by the medical investigator; pregnant (as evidenced by a positive pregnancy test) or nursing women; or subjects with medical or physical contradictions to the exercise intervention as determined by the medical investigator.

**Screening**

Screening Visit 1:As part of the first screening visit, each participant reviewed the Informed Consent with a member of the Outpatient Clinic staff. Once the consent was signed, the following was completed: height, weight, and vital signs; circumferences (waist, hip, and neck); medical history and physical exam; blood work, including lipid panels and liver function tests; women had a urine pregnancy test; resting 12-lead ECG; and questionnaires (Minnesota Leisure Time Activity and 3-day food record instructions were provided).

Screening Visit 2: On the second screening day, the participant reported to the Inpatient Unit to complete a 75g oral glucose tolerance test (OGTT). Blood was collected at specified time points up to 180 minutes post-glucose load, and urine was collected from the start of the OGTT until the 120-minute post-glucose load time point. After completing the OGTT, the participant was instructed to maintain current dietary patterns and not initiate a new exercise program. A lifestyle consultation was completed to assess the participant's readiness to initiate a physical activity program and assess barriers to participation. Patients taking medications for T2D were asked to withhold for at least 48 hours prior to the OGTT. Glucose was evaluated in the evening prior to and the morning of the OGTT. Testing did not proceed and was rescheduled if concentrations >300 or <50 mg/dL were observed.

**Diet/Weight Stabilization Period and Program Testing**

Participants were required to be weight and diet stable leading into test periods. They had to be stable for at least one week, but timing was adjusted if needed for stabilization. Once it was determined by study staff that the participant was stable and eligible for admission, the participant was admitted to the inpatient unit for testing. Patients taking medications for T2D were asked to withhold for at least 48 hours prior to the euglycemic clamp test (i.e., the morning of Admit Visit 1 and 4). Glucose was evaluated in the evening prior to and the morning of the clamp test. Testing did not proceed and was rescheduled if concentrations >300 or <50 mg/dL were observed.

Note: Females had to have their inpatient testing periods completed during the mid-follicular phase of the menstrual cycle. Therefore, the timing of the testing period from screening to the first test period and end of exercise and the second testing period was longer if necessary for scheduling purposes. Stabilization was assured as described above.

**Admit Visits**

<u>Admit Visit 1</u>: The participant arrived at the inpatient unit of PBRC in the late afternoon for an overnight stay for dietary control. Weights and vital signs were collected, along with completed 3-day food records. Once completed, the participant was provided dinner prior to an overnight fast and was allowed to sleep.

Admit Visit 2: The participant completed a DXA Scan in the morning. Females had a urine pregnancy test prior to the scan. An Aerobic Capacity Test was performed in the Exercise Testing Lab. The participant was allowed to leave the center when these procedures were completed, following the established weight/activity stabilization program. The participant returned to the inpatient unit at approximately 5 PM for an overnight stay for dietary control. Dinner was provided prior to an overnight fast.

Admit Visit 3: The participant underwent a Euglycemic Clamp Test. Metabolic rate was measured by RMR procedures four times (upon waking and prior to, during, and at the end of the clamp procedure), and two muscle biopsies were completed in conjunction with the clamp procedures. Participants were instructed to keep a 3-day food record prior to the next inpatient testing period. A lifestyle consultation was completed to discuss randomization group assignment and study expectations. Upon completion of the clamp procedure, a meal was provided, and the participant was discharged from the inpatient unit.

**Randomization**

Participants were randomized in blocks of 4 in a 1:1 ratio into one of the two treatment arms. The biostatistician provided sealed and sequentially numbered envelopes containing the treatment assignment to an unblinded member of the clinical research staff to notify the participant of their assignment. Envelopes were selected in sequence and were not opened prior to randomization.

**Interventions**

Exercise Group: All exercise sessions were supervised by the Interventional Resources Team and were conducted either in the Fitness Center at Pennington Biomedical Research Center or at an off-campus exercise environment approved by the study team. Alterations to the study intervention were made on a case-by-case basis as approved by the study team. Exercise consisted of walking/running on a treadmill and stationary cycling on a cycle ergometer. Subjects trained 5 days/week, approximately 60-90 minutes per session for 12 weeks. Initially, exercise was prescribed at 55-60% of heart rate max ($HR_{MAX}$) and was gradually increased so that after 2-3 weeks, subjects were exercising at 80-85% HRMAX (~70% $VO_{2MAX}$).

Control Group: Patients randomized to control were provided with current ACSM guidelines on exercise and physical activity, as well as dietary recommendations in accordance with the 2015-2020 USDA guidelines.

**Post-Intervention Testing**

Upon completion of the 12-week intervention, the participant was re-admitted to the inpatient unit for three days.

Admit Visit 4: The participant arrived at the inpatient unit of PBRC in the late afternoon. Weights and vital signs were collected, along with completed 3-day food records. Once completed, the participant was provided dinner and went to to sleep.

Admit Visit 5: The participant completed a DXA Scan. Females had a urine pregnancy test prior to the scan. An Aerobic Capacity Test was performed in the Exercise Testing Lab. The participant was allowed to leave the center when these procedures were completed and returned to the inpatient unit at approximately 5 PM for an overnight stay. Dinner was provided.

Admit Visit 6: The participant underwent a Euglycemic Clamp Test. Metabolic rate was measured by RMR procedures three times (prior to, at the beginning, and at the end of the clamp procedure), and two muscle biopsies were completed. Upon completion, a meal was provided, and the participant was discharged from the inpatient unit.

**Supplementary Methods 3**

**Skeletal Muscle Lipid Droplet and Contractile Structure**

~25 mg of muscle tissue was collected at 4% paraformaldehyde, 2% glutaraldehyde in 0.1 M phosphate buffer, pH 7.2 and stored at -4°C until time of assay. Samples were cut into 1 mm sections and placed in a 2.5% glutaraldehyde, 1% paraformaldehyde and 0.12 M sodium cacodylate solution, pH 7.2 overnight at room temperature. The following day, samples were washed five times at 3 minutes per wash with 0.1 M cacodylate buffer at room temperature. Samples were then placed in a 4% osmium solution (3% potassium ferrocyanide, 0.2 M cacodylate, and 4% osmium) for 1 hour on ice. Washes were performed after the osmium staining five times, 3 minutes per wash, with double distilled water. The samples were incubated again with a thiocarbohydrazide solution for 20 minutes at room temperature. Samples were stained with 2% osmium again for 30 minutes on ice and then washed again (five washes, three minutes each) with double distilled water. The samples were finally incubated overnight in a 1% uranyl acetate solution at 4°C. Half a liter of water was placed at 20°C overnight. The next day began by washing the samples five times, 3 minutes per wash, with the warmed double distilled water. The samples were placed in a Walton's lead aspartate solution (0.02 M lead nitrate and 0.03 M aspartic acid, pH 5.5) for 20 minutes at 20°C. After the incubation, samples were washed with room temperature double distilled water. Samples were dehydrated with an increasing concentration of ethanol washes (20%, 50%, 70%, 90%, 95%, and 100%; 5 minutes each) and then incubated in a 50% Epon: 50% ethanol solution for 4 hours in a vacuum sealed container. The last incubation of the day was performed by placing the samples in a 75% Epon, 25% ethanol solution overnight at room temperature in a vacuum sealed container. The last day of sample preparation consisted of three 100% Epon resin incubations for 1 hour, 1 hour, and 4 hours, at room temperature. Samples were place onto aluminum SEM mounts (Carl Zeiss Microscopy GmbH, Jena, Germany) after removing excess resin and placed in a 60°C oven for 48 hours to polymerize. After polymerization, the sample stubs were mounted in a UCT Ultramicrotome (Leica Microsystems, Inc., Deerfield, IL, USA) and faced with a Trimtool 45 diamond knife (DiATOME, Nidau, Switzerland) at a thickness feed of 100 nm and a rate of 80 mm/s. Images were also collected using an in-column energy selective backscatter with a filtering grid to reject unwanted secondary electrons and backscatter electrons, up to a voltage of 1.5 kV with a working distance of 5.01 nm. The milling function was performed with a FIB operating at 30 kV and a 2 – 2.5 nA beam current. The thickness of the FIB slices are 10 nm, providing a total volume thickness of 13–20 μm. Lastly, image stacks were aligned with a proprietary algorithm by using the Atlas 5 software and exporting the images at a TIFF file for future analyses. Lipid droplets were segmented using the deep learning model called MitoNet.[48] Using the high-performance computing system called BioWulf (NIH, Bethesda, MD), the image stacks were loaded into the Napari software. MitoNet can be accessed with the Empanada-Natpari plugin. Within the empanada plug in, training models were created and updated for each sample (Getting Started — empanada-napari-v1.1.1 0.1.1 documentation). Trainings were performed in the XY, XZ, and YZ planes of the images. Once an acceptable model was created, 3D inference was performed using a 0.5 segmentation confidence threshold, and ortho-plane parameters selected. The predicted labels were then saved as a Tiff file for further lipid droplet analysis. Myofibrils were segmented as described previously.[61] Briefly, raw FIB-SEM image volumes were rotated so that the XY images were of the muscle cell cross-section. The cross-sectional shape of sarcomeres in the first image of the volume were traced manually and then additional sarcomere cross-section tracing was performed sequentially at H-zones, Z-disks, and myofibril branch points for each connected sarcomere in series using interpolation between tracings. At sarcomere branch points, the previous tracing was continued along one part of the branch, while a new tracing was created for the other part allowing for multi-color representation of each myofibrillar segment within the connected myofibrillar matrix. Accuracy of segmentations was assessed by overlaying the segmented structures on the raw image files to ensure no errors in connectivity or sarcomere structure. The resulting Tiff file with the lipid droplet segmentation labels were loaded into ImageJ (NIH, Bethesda, MD). The image stack was duplicated, and a binary threshold was applied so that all the labels were assigned a 255 value, and the background was 0. The histogram function was performed on the whole image stack to get the total amount of pixels that made up the lipid droplet volume (all the pixels measured at 255) per total pixels that made up the image stack. Individual lipid droplet analysis was performed on the original lipid droplet segmentation labels. Using the MorphoLibJ plug-in in ImageJ, measures of voxels, volume, surface area, and sphericity were made using “Analyze 3D”. To measure lipid droplet length, the “Geodesic Diameter 3D” tool found within the MorphoLibJ plug-in was used.
